# Machine Learning-Driven Classification of Type 2 Diabetes Using Gut Microbiome Profiles for Enhanced Detection and Personalised Therapeutics

**DOI:** 10.64898/2025.11.26.25340813

**Authors:** Ayan Dharod, Tara Pratapa

## Abstract

The purpose of the study is to investigate Type 2 Diabetes Mellitus (T2DM) progression and risk through investigating gut microbiome biomarkers, providing insights into disease status and potential synbiotic and fecal microbiota transplantation interventions. Gut dysbiosis serves as a key indicator for T2DM detection, supporting effective intervention strategies by outlining potential targets for up/down-regulation. Our preliminary research aimed to compile gut microbiome biomarkers documented to be associated with disease states, forming the foundation for constructing a model capable of disease state analysis. Gut microbiome composition data of individual patients from sequenced metagenomic files were compiled, encompassing the abundance of over 4,500 microbial taxa across profiles. Multiple models were tested to evaluate the greatest efficacy given the nature of the data used, from which XGBoost was selected and optimized. Despite 95.8% sparsity in our microbiome data alongside high local intrinsic dimensionality (LID≈12), an 80% testing accuracy was achieved. SHAP values enabled the analysis of key bacterial features, alongside a correlational network for biological validation through the visualization of co-occurring T2DM-associated bacterial taxa. This supported the hypothesis that machine learning can provide critical insights into the nature of dysbiosis, precisely classifying disease states and modelling biologically-accurate relationships. Our findings demonstrate the biological accuracy of the model, along with its ability to identify under-researched species significantly correlated with disease states. This suggested their potential as stable and reliable biomarkers. While further data is required to enhance model performance, this work represents a crucial step toward revolutionizing targeted diagnosis, patient care, and intervention strategies.

## 1. Introduction

Type 2 Diabetes Mellitus (T2DM) is a metabolic disorder characterized by the excess production of glucose, caused by the inadequate secretion of insulin by Beta-cells, resulting in hyperglycemic states^1^. Data from the 2022 NCD Risk Factor Collaboration reports that approximately 828 million individuals live with diabetes globally, with over 95% having type 2 diabetes mellitus. Type 2 diabetes not only affects older populations but has seen a two- to threefold increase among individuals below the age of 40^2^. Even more concerning is the fact that almost one in two adults with diabetes are undiagnosed, remaining unaware that they suffer from the disease^3^. This reflects a concerning gap in access to timely diagnosis and thereby enabling early intervention and treatment for the disease, despite its worldwide prevalence.

With millions affected and at risk of severe complications, patients would greatly benefit from the application of precision medicine that accounts for the inherently multifactorial nature of this disease^4^. For example, clinical trials reveal higher remission rates for T2DM patients taking personalised dietary interventions compared to generalised nutrition advice, highlighting the effectiveness of customized strategies to combat T2DM^5^. The employment of other such personalised therapeutic avenues, including fecal microbiota transplantation (FMT) and synbiotics, display great efficacy in improving patient health outcomes over time influencing microbial diversity ^6^.

Gut dysbiosis is a notable marker for the T2DM disease state. The human gut contains a diverse range of bacterial taxa, and research has demonstrated its key role in metabolic homeostasis. Disruptions in this balanced composition have the potential to drive progression of various diseases, including metabolic syndrome disorders like T2DM^7^. The availability of large datasets enables machine learning algorithms to identify key patterns in microbiome compositions of patient samples in the same group. This makes computational methods well suited for processing the heterogeneity of gut microbiome data, through the employment of techniques like feature identification, selection, and post-intervention strategy optimization ^8^.

The scope of the application of machine learning algorithms to handle diverse gut microbiome datasets led to the investigation of its efficacy in analysing disease state and suggesting intervention strategies based on this analysis. This project examines the gut microbiome as a source of biomarkers for detecting T2DM, and leverages computational approaches to aid its early detection and intervention. The main objective is improving methods for assessing a person’s risk of diabetes through biomarkers in the gut, further enabling personalized interventions for the given patient(s).

## 2. Methodology

### 2.1 Data Collection

Publicly available human gut microbiome datasets containing both healthy and T2DM cohorts were identified through a comprehensive literature search. Studies were included only if they provided relative abundance data from stool-derived bacterial species based on sequencing analyses. In order to ensure proper standardization of data, a clearly defined inclusion and exclusion criteria were constructed, as outlined in Table 1.

**Table 1:**
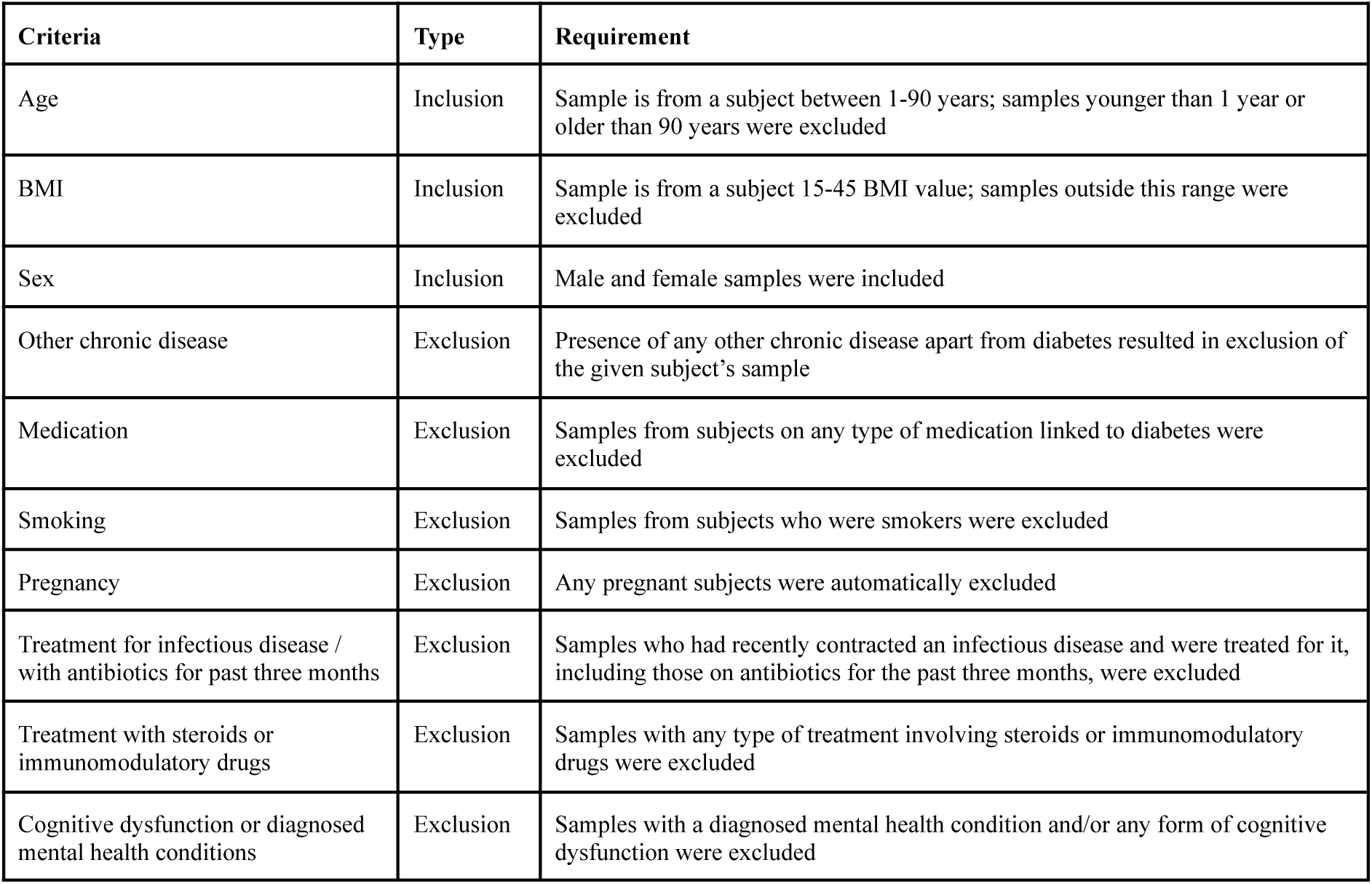
Inclusion and exclusion criteria to filter data samples from studies.

| Criteria | Type | Requirement |
| --- | --- | --- |
| Age | Inclusion | Sample is from a subject between 1-90 years; samples younger than 1 year or older than 90 years were excluded |
| BMI | Inclusion | Sample is from a subject 15-45 BMI value; samples outside this range were excluded |
| Sex | Inclusion | Male and female samples were included |
| Other chronic disease | Exclusion | Presence of any other chronic disease apart from diabetes resulted in exclusion of the given subject's sample |
| Medication | Exclusion | Samples from subjects on any type of medication linked to diabetes were excluded |
| Smoking | Exclusion | Samples from subjects who were smokers were excluded |
| Pregnancy | Exclusion | Any pregnant subjects were automatically excluded |
| Treatment for infectious disease / with antibiotics for past three months | Exclusion | Samples who had recently contracted an infectious disease and were treated for it, including those on antibiotics for the past three months, were excluded |
| Treatment with steroids or immunomodulatory drugs | Exclusion | Samples with any type of treatment involving steroids or immunomodulatory drugs were excluded |
| Cognitive dysfunction or diagnosed mental health conditions | Exclusion | Samples with a diagnosed mental health condition and/or any form of cognitive dysfunction were excluded |

Datasets were further collected ensuring proper diversity of various demographic factors. These included, but were not limited to, age, sex, ethnicity, geographic, and sociodemographic factors. Ensuring samples ranged from a diverse set of such factors ensured a balanced representation of the human population, ensuring the results and findings from this research were applicable to a wider set of individuals.

Two primary sequencing methodologies were used across selected datasets: whole-genome shotgun (WGS) metagenomics and 16S rRNA amplicon sequencing. WGS profiles all microbial DNA present in a stool sample, enabling species- and genus-level resolution^9,10^. Post-DNA extraction^11^, libraries were prepared and barcoded for multiplexed sequencing on Illumina platforms^12,13^. Host reads were removed using Bowtie2 ^14^, and species abundances were derived via MetaPhlAn^15,16^, producing final sample-level abundance tables. 16S rRNA amplicon sequencing targets the bacterial 16S gene to infer community composition at the species level^17,18^. In such workflows, the 16S rRNA gene was amplified by PCR^19^, followed by barcoded Illumina sequencing^20^. Reads were quality-filtered using DADA2 and Deblur^21^, demultiplexed^22^, and taxonomically assigned using the SILVA reference database to generate amplicon sequence variants (ASVs) and abundance matrices^23^.

Through these processes, research studies were able to curate relative abundance values for each bacterial species in each sample. Table 2 summarises the final research studies selected for data collection, including key features of the studies that render them applicable for this research.

**Table 2:** Summary of research studies used for data collection, and their features.

| Study ID | Method | Instrument | Valid Runs | Reads across Sample Runs |  |  |
| --- | --- | --- | --- | --- | --- | --- |
|  |  |  |  | Range | Mean | Median |
| PRJEB1786 | Metagenomics (WGS) | Illumina HiSeq 2000 | 147 | 3967737-73997021 | 15731843 | 13969150 |
| PRJNA289586 | Metagenomics (WGS) | Illumina HiSeq 2000 | 133 | 9678285-62783024 | 42581437 | 50349366 |
| PRJNA299502 | Metagenomics (WGS) | Illumina HiSeq 2000 | 19 | 13196473-30829874 | 23269051 | 23171029 |
| PRJNA389481 | Amplicon (16S rRNA) | Illumina MiSeq | 66 | 22929-51257 | 34644 | 33278 |
| PRJNA422434 | Metagenomics (WGS) | Illumina Genome Analyzer II | 359 | 6987735-49892410 | 20374459 | 18680470 |
| PRJNA448494 | Amplicon (16S rRNA) | Illumina HiSeq 2500 | 112 | 254741-1548277 | 641039 | 552490 |

### 2.2 Data Pre-Processing

All individual sample reads were compiled into one larger spreadsheet that would act as the parent dataset for the purpose of this research, from which an extract is shown as reference in Table 3. Here, columns represented bacterial species, while rows represented samples. A given abundance value could be understood based on the column it belonged to (which bacterial species the abundance was of) and the row it was present in (which species had this abundance level). A ‘0’ abundance value was inputted for any bacterial species not found in a sample’s gut microbiome. Finally, binary encoding was done for each sample based on the class they belong to: ‘0’ (healthy) and ‘1’ (diabetic).

**Table 3:**
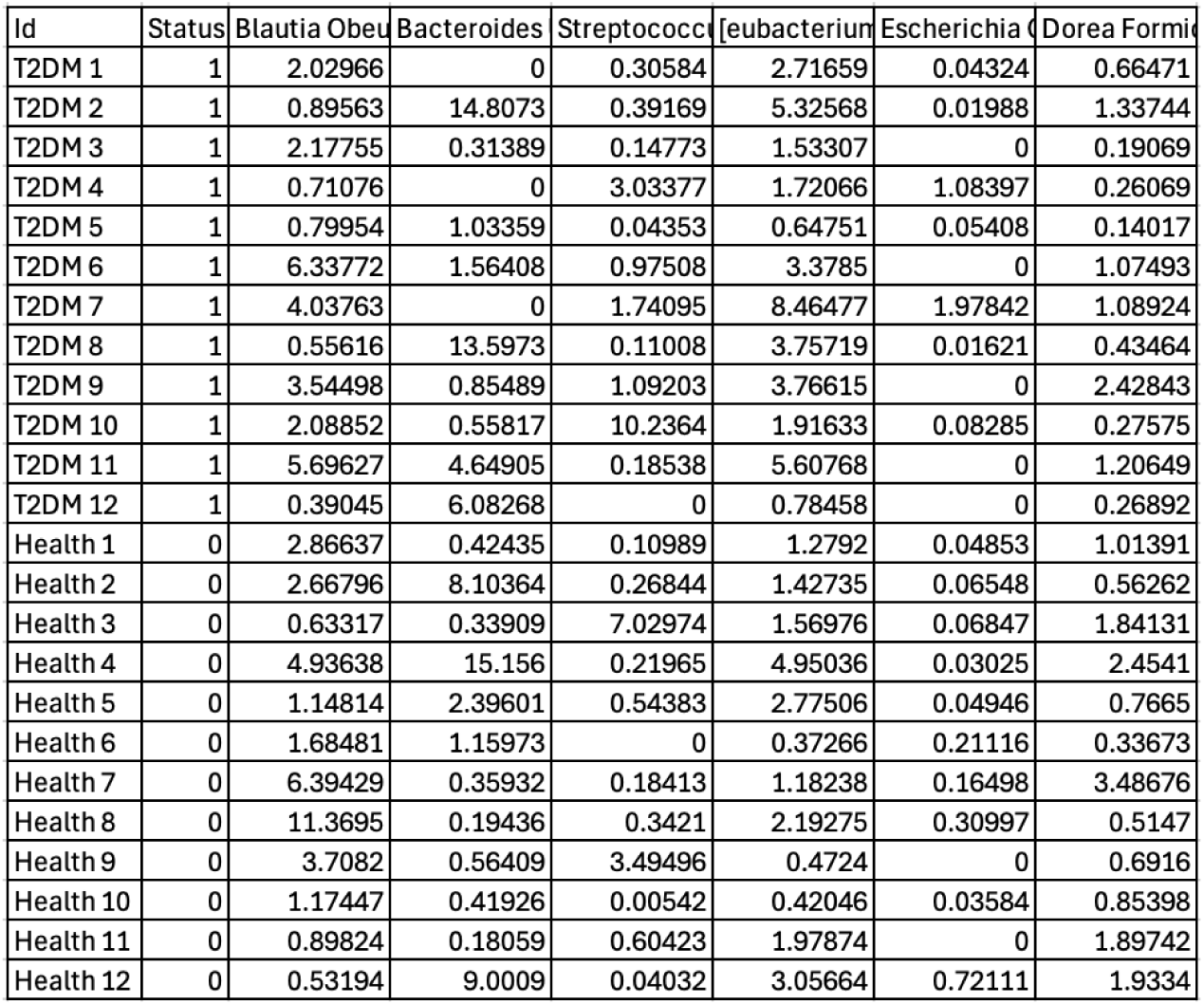
Excerpt of bacterial species abundance data for healthy and diabetic classes.

Quality control was performed to ensure data integrity, minimising the risk of misleading biological interpretations or false associations between the features and the target disease. Samples lacking critical metadata items (disease group, age, sex, BMI, nationality and presence of other disease conditions) were removed. Unassigned or unclassified bacterial species were also filtered out. A separation was made between true zeroes and unmeasured values; true zeroes were retained while unmeasured values were removed. Samples with >10% of bacterial species not measured were excluded. Additionally, those bacterial species present in <5% of samples were eliminated, reducing noise and overfitting. Missing abundance values were imputed by taking the median abundance of each bacterial species within the given group. Median was the preferred measure of central tendency given its robustness to outliers. Imputed values were flagged for later identification, and a final filtration was done of any sample or feature with over 20% imputed entries. This elimination ensured minimal fabricated data, preserving biological validity and natural variation.

Post-data cleaning, the final pre-processing steps were performed to eliminate any remaining discrepancies. Feature harmonization helped resolve duplicate taxa created by inconsistent naming across studies. Once complete, a sample-by-feature matrix was generated, with the remaining metadata stored separately. A train-test split was done in the 80:20 ratio respectively. Class balance was ensured across splits to prevent model bias towards any majority class.

### 2.3 Constructing the Machine Learning Model

Binary class labels were assigned to each outcome, after which a random forest (RF) model was trained on the dataset. RF was selected due to the data’s high dimensionality and sparsity, with features greatly exceeding the number of patient samples. Such a model can mitigate the overfitting commonly seen in single decision trees trained on the given biological data’s nature. As an ensemble method, it reduces variance by averaging predictions from multiple, diverse trees and thereby reducing model variance^24^.

**Figure 1:**
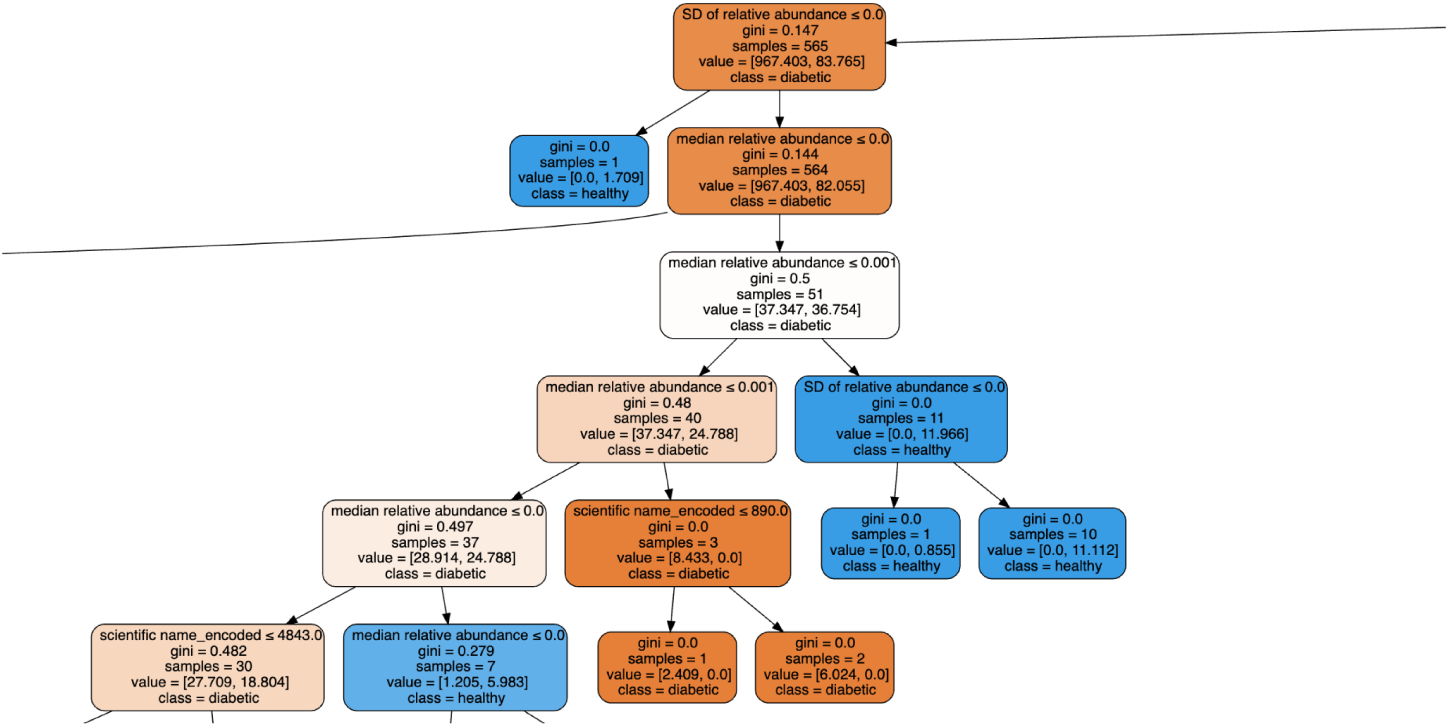
Excerpt of a decision tree within the Random Forest model.

The implemented random forest model yielded an accuracy of 0.82. However, there was significant overfitting and low biological validity, as found through discrepancies between top SHAP-derived feature importances and expected biological relevance. The features used to train the RF model include: number of samples in which a species was detected, mean relative abundance, median abundance, and standard deviation of abundance. The data’s high sparsity, with many zero and near-zero values, combined with high dimensionality, worsened model overfitting. Too many predictors relative to samples exacerbated this. Derived statistical measures such as standard deviation amplified noise, which the random forest could not fully mitigate. While RF constructs each tree on a random subset of features and samples, each tree can individually overfit noisy, high-dimensional input which was present in the dataset, and generalisation was impaired.

After evaluating the RF model, simpler regression-based approaches were tested to compare performance and interpretability. Recognizing that training off of derived statistics (mean, median, and standard deviation) increased input noise and overfitting, subsequent models were built using solely abundance values to improve signal-to-noise ratio and biological interpretability. XGBoost, LASSO, Elastic Net, and Ensemble regression models were evaluated. Each regression model was selected to enhance generalizability by learning from abundance features across patients while managing sparse data through selective feature weighting. This balanced bias and variance control for improved stability and interpretability of features. LASSO and Elastic Net applied built-in regularization constraints to minimize overfitting in such high-dimensional data. The ensemble regression model was implemented to combine multiple weak learners, enhancing predictive robustness while also reducing variance. Performance results of the models based on standard ML metrics are shown in Figure 2.

**Figure 2:**
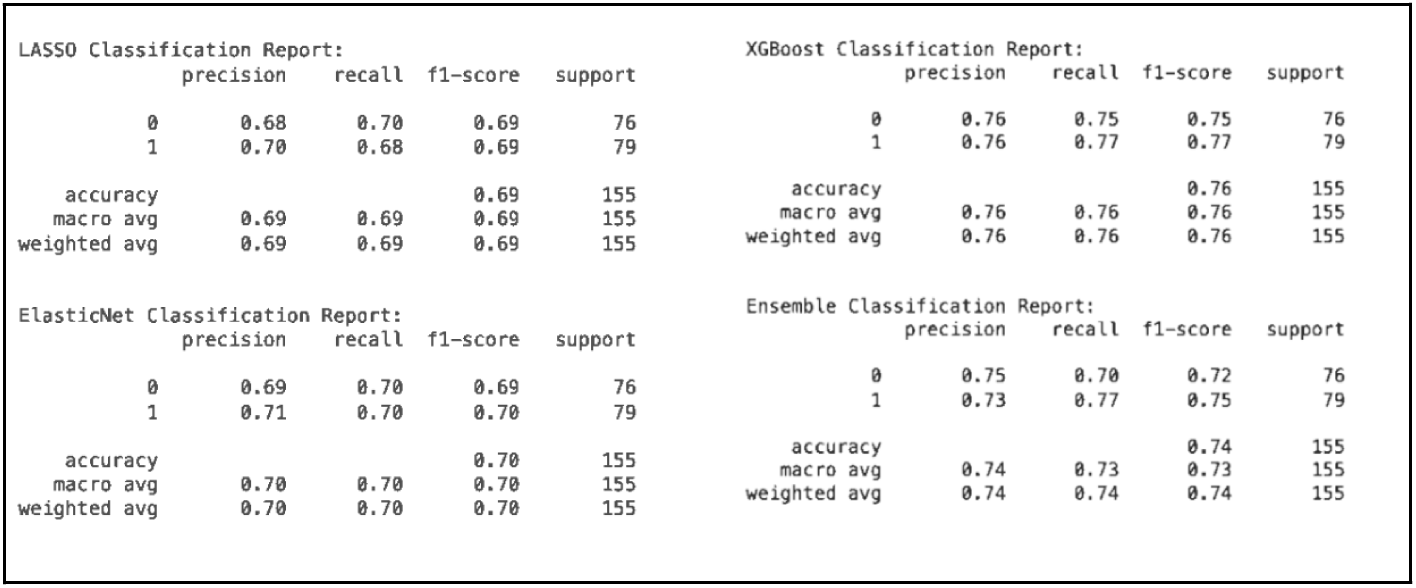
Performance comparison across regression-based models.

XGBoost, a gradient-boosted ensemble, was specifically selected because of its ability to sequentially correct prior errors and natively handle sparse data, addressing both bias and variance. The model’s sequential tree-building process improved generalization over RF, improving the biological plausibility of selected features. XGBoost achieved the best performance compared to the other models with a testing accuracy of 0.7613. Its iterative error correction and regularized approach sequentially corrects prior mistakes, reduces model complexity, and accommodates sparsity with built-in mechanisms.

Only 458 columns contained >10% non-zero values, prompting tests of higher density thresholds, where datasets were reconstructed using limits to filter features with >30% and >50% non-zero values. However, model accuracies decreased when trained on these filtered datasets. This approach inadvertently excluded biologically important yet sparse features that showed significant variation between diabetic and healthy groups. The omission of these features also degraded SHAP-reported feature importance profiles, ultimately reducing biological accuracy.

Age and BMI values for each patient were collected but had not been incorporated into the model initially. These variables were subsequently included, as they were hypothesized to be indicative of metabolic status and potentially improve generalization by providing physiologically relevant context. However, with a significant amount of BMI data available only as categorical ranges rather than precise values, its inclusion led to a slight reduction in model accuracy, suggesting limited predictive benefit under the current data conditions. Therefore, the final model only included age as a demographic feature from sample metadata.

**Table 4:** Precision and Recall Analysis Post–Dataset Alterations.

| Dataset | Precision (0, 1) | Recall (0, 1) |
| --- | --- | --- |
| *Original* | 0.76, 0.76 | 0.75, 0.77 |
| *30 percent (Columns)* | 0.73, 0.72 | 0.70, 0.75 |
| *50 percent (Columns)* | 0.75, 0.73 | 0.71, 0.77 |
| *With age and BMI* | 0.73, 0.71 | 0.68, 0.76 |

When calculating SHAP values, there remained a lack of biological accuracy and scope for improving generalization through refined feature selection. To address this, two complementary approaches were implemented: Gradient Boosting Machine (GBM) and SelectKBest using *f_classif* for feature selection. Since the model continued to demonstrate overfitting, these methods were applied to ensure that the selected features enhanced generalizability. Applying both techniques helped mitigate the effects of high dimensionality by ensuring that trees utilized consistently differentiating or statistically significant features for prediction, thereby improving model performance.

GBM provided a measure of feature importance, quantifying the contribution of each feature to model accuracy during training. SelectKBest ranked features based on the ANOVA F-test for classification, selecting the top K features with the highest scores. Following implementation and SHAP-based feature evaluation, there was a notable increase in biological accuracy. More of the identified species corresponded to documented gut microbiome biomarkers associated with diabetes, accompanied by a measurable improvement in predictive accuracy. Among the top 15 SHAP values, 7 overlapping features were observed between the GBM and SelectKBest results, reinforcing the robustness of the final feature set.

In order to identify the optimal combination of parameters that would support the greatest model performance, GridSearchCV was used. Grid search systematically evaluates a predefined dictionary of hyperparameter values, forming a grid of all possible combinations and iteratively testing each one^25^. The k-fold cross-validation method was conducted on each combination, enabling the identification of the best performing hyperparameters. Here, the training data was split into 5 (value of ‘k’) folds, with the model being trained on 4 (value of ‘k-1’) folds. The remaining fold was used to validate the performance of the model for the given combination, calculating accuracy as the key metric, and the process was repeated 5 (‘k’) times to calculate the average validation score across folds^26^. After conducting k-fold cross-validation across all combinations, the parameter set with the best average score was chosen as the optimal model hyperparameters. Considering the fact that each combination of hyperparameters is evaluated using k-fold validation, each fold serves as a temporary validation set for hyperparameter analysis but is not involved in tuning for other folds. Selection reflects performance on unseen data splits. Furthermore, choosing hyperparameters that result in the best validation scores means the model is generalizing across varying data. For GBM, changes including adjusting the number of estimators (129) and maximum depth (4), and refining ultra-sparse feature thresholds increased model accuracy from 0.73 to 0.78. For SelectKBest, changes including tuning the number of selected features (K=1480) and updating the sparsity threshold improved accuracy from 0.76 to 0.80. These parameter refinements ensured more stable feature selection and better representation of biologically relevant features.

### 2.4 Interventions

#### 2.4.1 Synbiotics

Probiotics contain live bacteria with beneficial effects, while prebiotics are nutrients that promote the proliferation of these beneficial microorganisms. Together, they synergistically aim to enhance the growth of targeted species like Bifidobacterium and Lactobacillus, improving gut health. In combination with the addition of substrates such as lactose, which support bacterial growth, they form a synbiotic. This combination results in a greater overall effect, specifically designed to support the newly introduced beneficial bacteria and improve gut microbiota balance^27^.

The analysis of patient-derived microbiome samples identified key microbial taxa contributing to gut dysbiosis and potential progression toward disease states. Quantitative assessment of compositional profiles provided insight into taxa whose relative abundances deviated significantly from those characteristic of a healthy microbiome. These deviations were used to inform targeted modulation strategies through the administration of synbiotics, designed to restore microbial equilibrium.

Taxa exhibiting abundances exceeding or falling below established healthy thresholds were prioritized for individualized intervention.

The magnitude of deviation from these thresholds informed the formulation of patient-specific synbiotic compositions, comprising agents that either promote or suppress the growth of particular bacterial taxa. This precision-guided framework enables a tailored therapeutic approach aimed at reestablishing microbial homeostasis and mitigating the functional consequences of gut dysbiosis.

**Figure 3:**
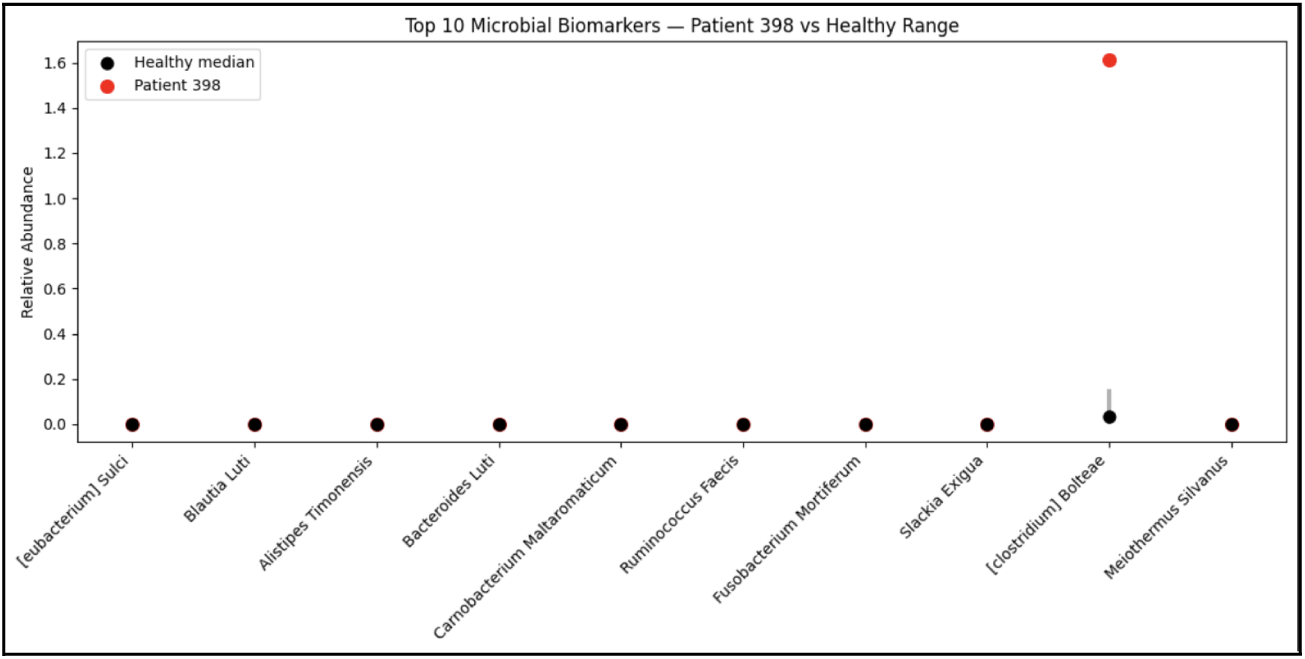
Comparative Abundance of Key Bacterial Taxa in Diabetic vs. Healthy Patients.

Individual patient analyses for specific microbiome features are illustrated in the figures. Key indicators of dysbiosis can be identified by examining bacterial taxa within patient samples that significantly contribute to the disease state. In the depiction above, diabetic patients exhibit species-level abundances that differ from the median values observed in healthy controls. These deviations can be mapped to bacterial species within a repository containing evidence-based synbiotics known to upregulate or downregulate specific taxa. In the first patient, targeted intervention may be beneficial due to the observation of elevated *Clostridium bolteae* levels. The synbiotic combination *Clostridium butyricum* with chitooligosaccharides (COS) has been tested and shown to inhibit pathogenic *Clostridium* species, improving intestinal barrier integrity and reducing inflammation^28^. This demonstrates how individualized microbiome analysis can be integrated with a synbiotic intervention repository to enable microbiome-targeted intervention.

#### 2.4.2 Fecal Microbiota Transplantation

Fecal microbiota transplantation (FMT) refers to the transfer of a healthy donor’s stool (believed to have a balanced gut microbiome) to a diseased recipient (with an unbalanced microbiome composition)^29^. The aim of this intervention is restoring the microbial balance of up- or down-regulated gut elements in the diseased recipient, reversing dysbiosis^30^. Healthy samples from the initial data sheet were taken as potential donors, with 50 diabetic samples randomly selected as recipients of FMT. During preprocessing, only bacterial species present in at least 10% of samples were retained to remove extremely rare features.

Across the healthy samples, key measures of central tendency were calculated, including mean, median, standard deviation, and the 10th and 90th percentiles. These became the benchmarks of each bacterial species as it would exist in a healthy, balanced gut microbiome, enabling comparison of excess and deficit species in the diabetic patient. For each Type 2 Diabetic patient (recipient), three key metrics were calculated: deviation from the mean, Z-score, and percentile position. Given the high variance in diabetic sample species abundance compared across the healthy donors, values of ‘0’ and ‘100’ were given if the values were lower than the minimum or exceeding the maximum respectively, handling outliers during percentile ranking.

Donor-recipient mapping began by identifying taxa in each diabetic patient requiring modulation, using Z-score and percentile ranks to classify over- or under-represented species. A conditional framework combined these criteria to label each taxon as high, low, or normal in abundance, prioritizing significant deviations as biologically relevant. Figure 4 highlights the step-by-step process through which a verdict was defined on whether modulation was needed for a given bacterial species.

**Figure 4:**
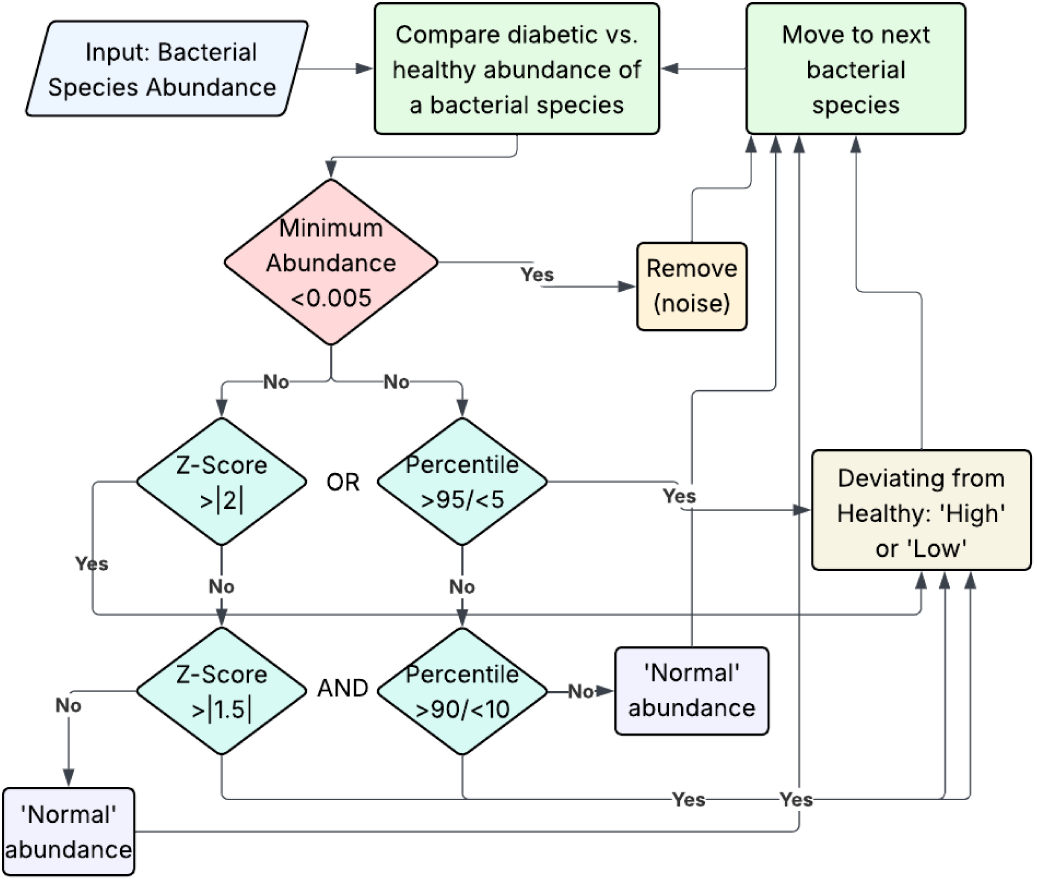
Flowchart showing evaluation of each bacterial species (need for regulation)

A donor-species matrix was crafted by calculating each taxon’s abundance across all healthy samples. percentile ranks for each bacterial species across potential donors. This helped identify the placement of different donors for a given bacterial species presence in context of the remaining samples, which would aid in ranking candidates for the given diabetic recipient. For each recipient, only dysregulated bacterial species were considered. Percentile values were converted into directional percentiles: higher percentiles favored donors capable of upregulating deficient taxa, while inverted percentiles favored donors suited to downregulate excessive ones.

These directional percentiles were weighted by the absolute value of the Z-score for each taxon, emphasising those species which were deviating more significantly. A weighted average was calculated to produce donor-specific compatibility scores, where increased scores indicated better donor-recipient matches. Accordingly, the weighted averages were ranked in descending order. The donor ID was retrieved for the corresponding score, enabling a final list of the top donors for a given recipient.

## 3. Results

### 3.1 Model Optimization and Performance

The final testing accuracy achieved for the model was 80.00%. Alongside accuracy, other metrics were also measured to evaluate the model’s performance, as shown in Table 5.

**Table 5:** Model metrics for evaluating performance and effectiveness.

| Class | Precision | Recall | F1-Score | Support |
| --- | --- | --- | --- | --- |
| Healthy ('0') | 0.80 | 0.79 | 0.79 | 76 |
| Diabetes ('1') | 0.80 | 0.81 | 0.81 | 79 |
| Average | 0.80 | 0.80 | 0.80 | 155 (sum) |

A confusion matrix of the XGBoost model predicted 64 true positives, 60 true negatives, 16 false positives and 15 false negatives, as detailed in Figure 5.

**Figure 5:**
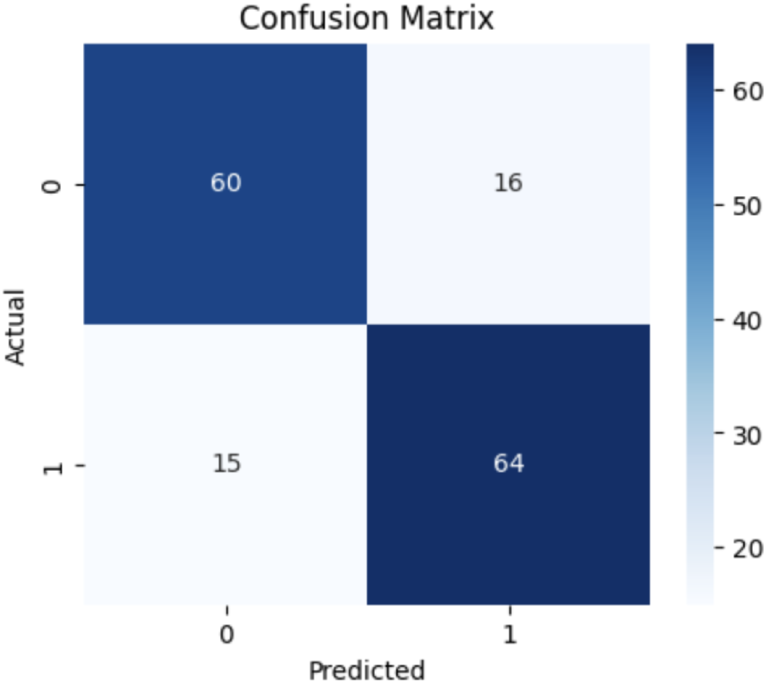
Confusion matrix for the XGBoost model.

Figure 6 depicts the ROC/AUC curve plotted for the XGBoost model. The blue line indicated the baseline on which a model is compared, acting as a random guesser with an AUC of 0.5. The XGBoost model achieved an AUC value of 0.856, indicating strong discriminative ability with high true-positive rates and low false-positive rates.

**Figure 6:**
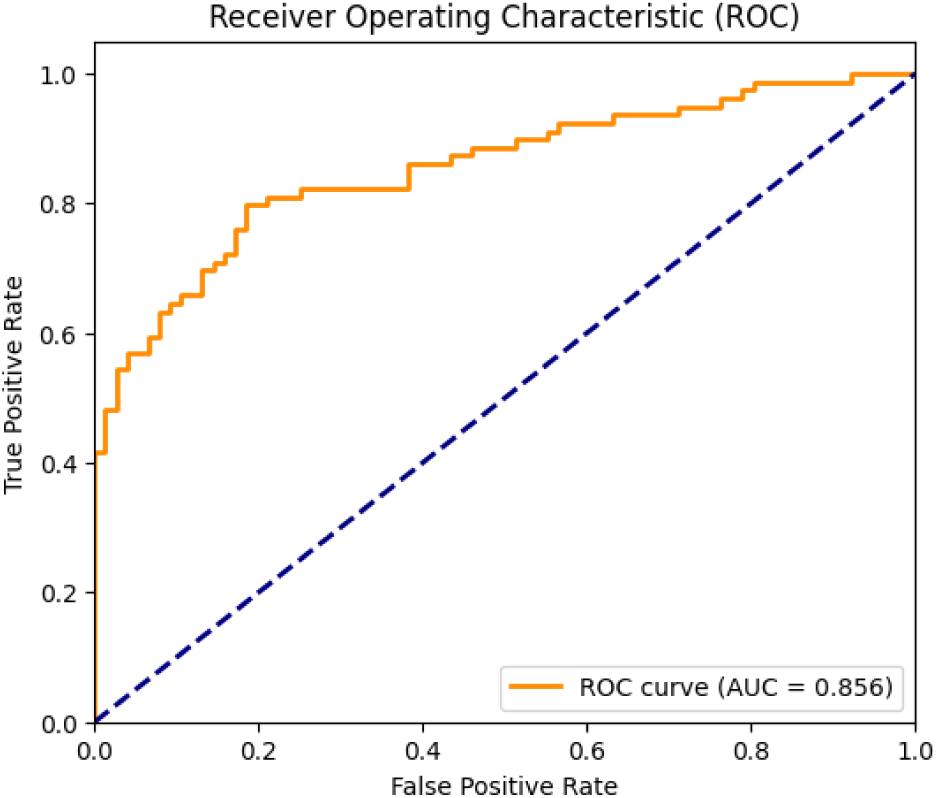
ROC/AUC curve for the XGBoost model, showing strong discriminative ability.

Figure 7 depicts the calibration curve plotted for the XGBoost model. The dashed grey line represents a perfectly calibrated model where the predicted frequency of positives is equal to the true frequency of positives. The constructed XGBoost model is compared with the dashed line, showing good calibration in the low (very likely healthy) and high (very likely diabetic) ranges, though it deviates more in the middle range for more borderline cases.

**Figure 7:**
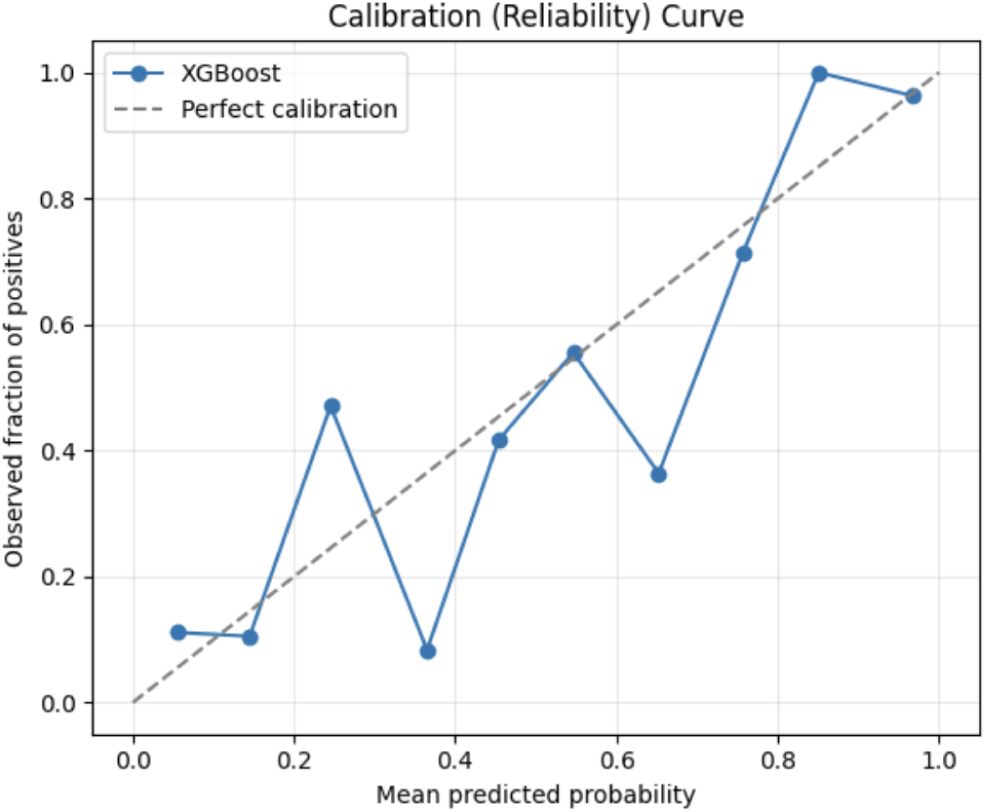
Calibration curve for the XGBoost model.

### 3.2 Regularization and Generalization Performance

In order to compare the training and testing accuracy of the model, two charts were created to highlight how the two accuracy values changed over boosting rounds. One of the charts was made based on an XGBoost model that employed feature selection, while the second was made with all the features, acting as a comparison to evaluate the effectiveness, or lack thereof, of feature selection in improving model outcomes (measured through accuracy).

Figures 8 and 9 represent the aforementioned charts with and without feature selection respectively. A comparison of the training and testing accuracy curves is critical in highlighting key aspects of the model. Both charts show an initial rise in accuracy, reflecting early model learning. The widening gap between training and testing curves indicates overfitting, where the model memorizes data patterns and achieves near-perfect training accuracy (1.00). Further, we see a plateauing of the testing accuracy curve after ≈40 boosting rounds, indicating diminishing returns from additional training. This highlights the models’ ability to capture most microbiome-related signals within 40-60 boosting rounds, beyond which additional trees primarily fit noise as training accuracy rises while generalizability declines. Hence, a key outcome of these two charts is understanding the optimum number of boosting rounds needed for the given data to train an XGBoost model optimally.

A comparison of the two charts highlights the importance of feature selection, compared to the inclusion of all features for the model. Figure 8 achieves a higher testing accuracy ceiling than Figure 8, demonstrating superior model efficacy when feature selection is employed. Figure 9 also illustrates a wider gap between train and test accuracies, indicating greater overfitting when all features are retained. Finally, greater stability is seen in Figure 8, highlighting fewer fluctuations with feature selection. Each of these comparisons accentuate the biological basis behind the importance of feature selection. Given the high dimensionality and sparsity of microbiome data, an inclusion of all features leads XGBoost to memorise training data and lose generalisation as a result. By removing irrelevant and redundant species, feature selection enables improved model performance while also maintaining stability and reducing overfitting.

**Figure 8:**
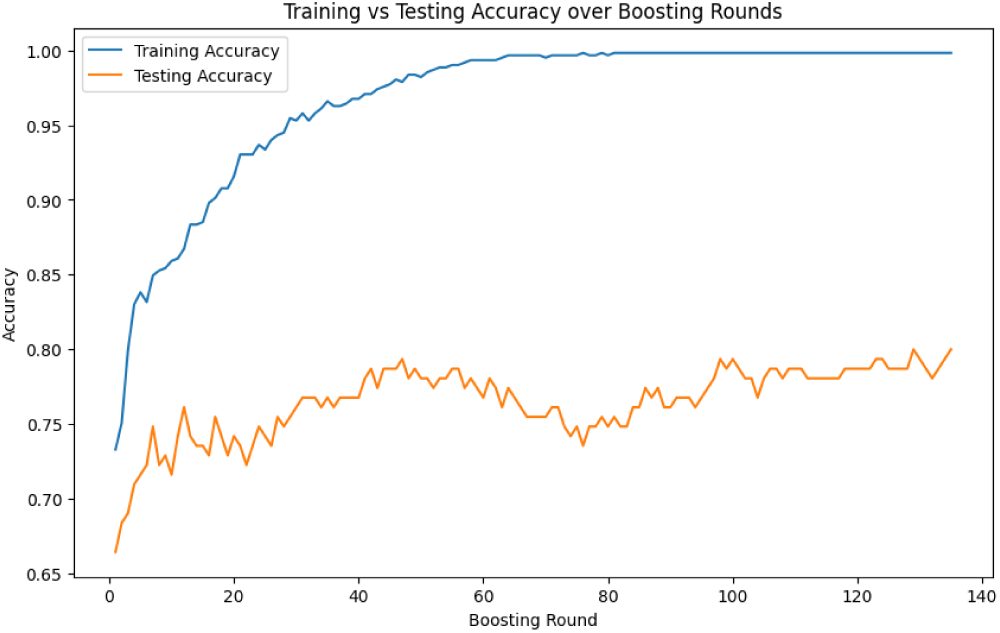
Training & testing accuracy over boosting rounds for XGBoost model (feature selection)

**Figure 9:**
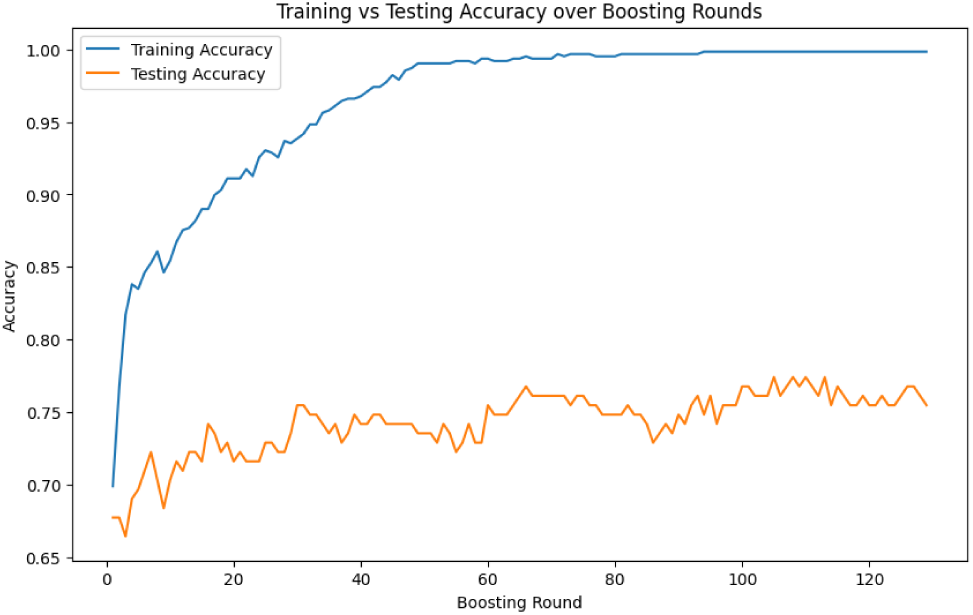
Training and testing accuracy over boosting rounds (without feature selection)

Building on the inferences made on the importance of feature selection, a chart was plotted to deduce the relationship between an increasing number of features (as described by ‘k’ values from the SelectKBest method of feature selection) and the resulting model accuracy. The chart was built post-filtering the ultra-sparse features, with <2000 features of the original ≈4500 features for the SelectKBest method. Iterations were conducted at a step value of 20, measuring the model accuracy with an increasing number of features. Figure 10 depicts this relationship. An evident finding from the chart is the absence of monotonic improvement of the model correlating with the number of features included. As can be seen, the highest accuracy value was achieved not with the complete set of non-ultra-sparse features, but in fact from a lower number of selected features (≈1440). This highlights how fewer features, in certain cases, aid in decluttering and simplifying data for a model to train on, enabling improved performance. Selecting only the most informative features enhances generalization and facilitates denoising of the data, since the inclusion of more redundant bacteria increases dimensionality while not adding signal. Therefore, alongside simply ultra-sparse feature elimination, purposeful feature selection using computationally-backed methods is critical when working with data of the nature of microbiome samples.

**Figure 10:**
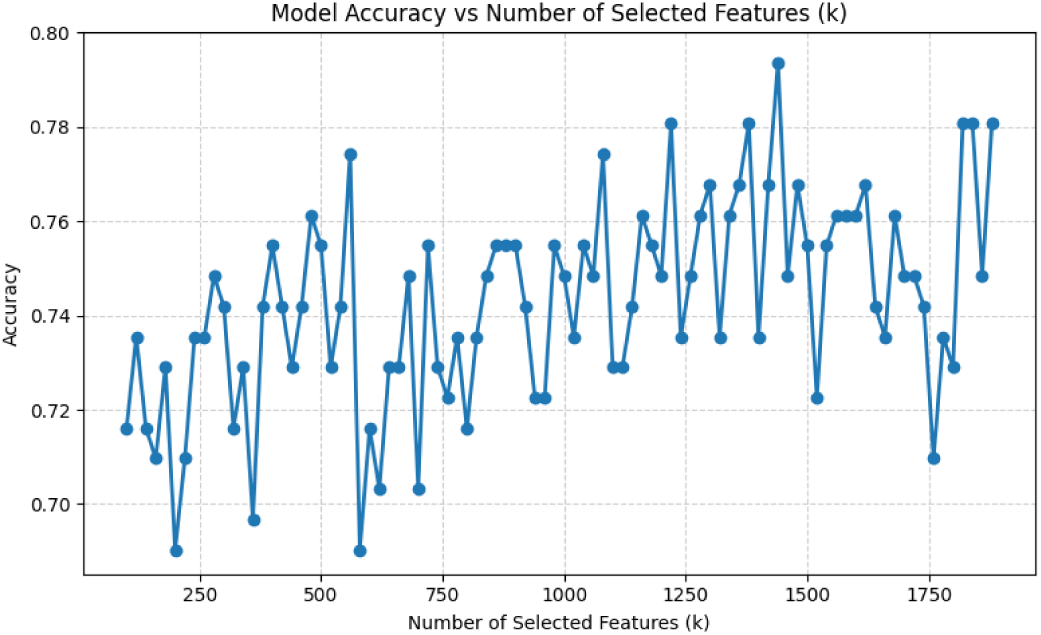
Model accuracy over across varying number of features selected (‘k’)

### 3.3 Model Explainability and Feature Robustness

SHAP values describe the magnitude of contribution of a given feature, with a positive or negative sign indicating a push towards or away from the positive (diabetic) class. This supports global interpretability for a model’s most significant features, alongside the directional effect of a given feature’s values on the predicted risk of both an individual sample and the overall model.

A global SHAP feature importance plot helps rank different features (bacterial species) by their mean SHAP contribution, enabling comparison of each feature’s overall influence irrespective of direction. This supports the identification of the most influential microbiome species, helping validate feature selection and interpretability. Figure 11 details such a plot for the XGBoost model. The presence of age as the feature with the largest contribution is consistent with the strong age-linked microbiome patterns expected in T2DM. Other gut microbial species shown have prior associations with key biological processes involved in diabetes, including metabolism, inflammation and insulin resistance, highlighting the biological plausibility of the signals identified by SHAP.

**Figure 11:**
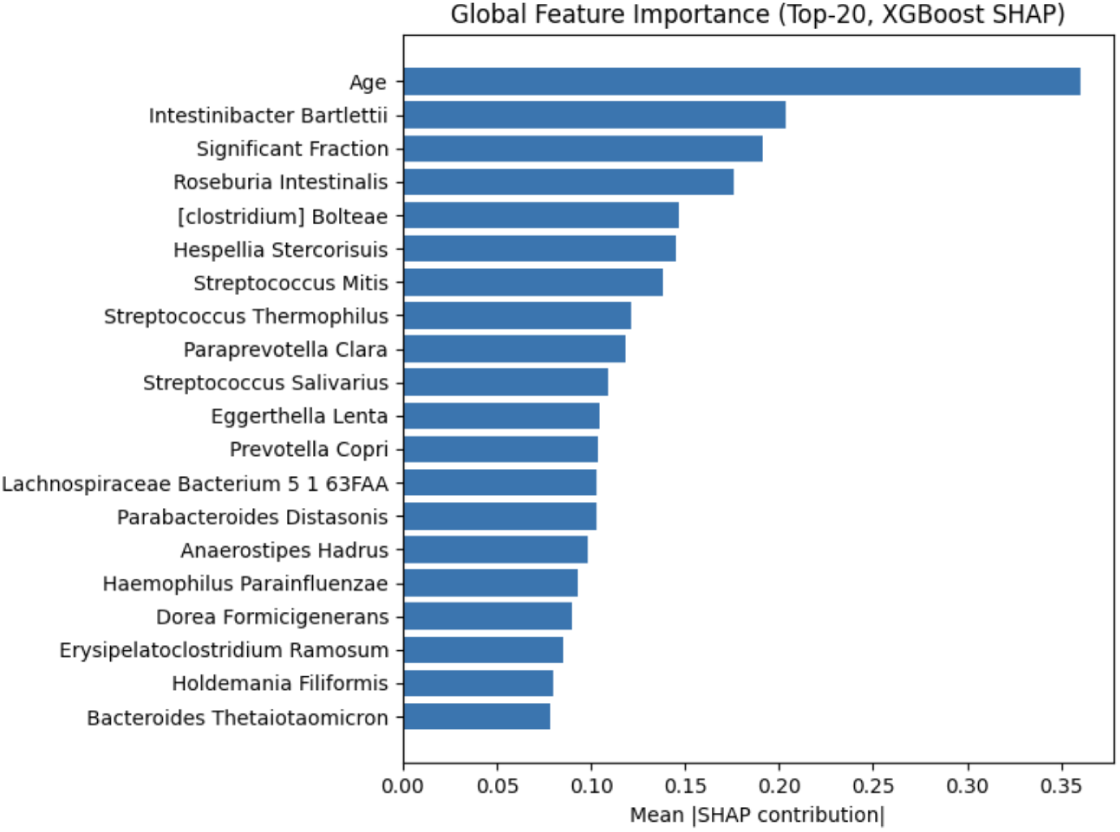
Global SHAP feature importance plot of the XGBoost model.

A beeswarm plot (Figure 12) visualises feature contributions at the individual-sample level for the top 20 bacterial taxa. Unlike the global SHAP importance plot, which reflects only magnitude, the beeswarm plot captures both the direction and magnitude of each feature’s effect. Each dot represents a sample’s SHAP value of a given feature, plotted against the x-axis, with color denoting feature abundance. A color gradient illustrates the influence of abundance on model prediction shifts towards or away from the diabetic class.

**Figure 12:**
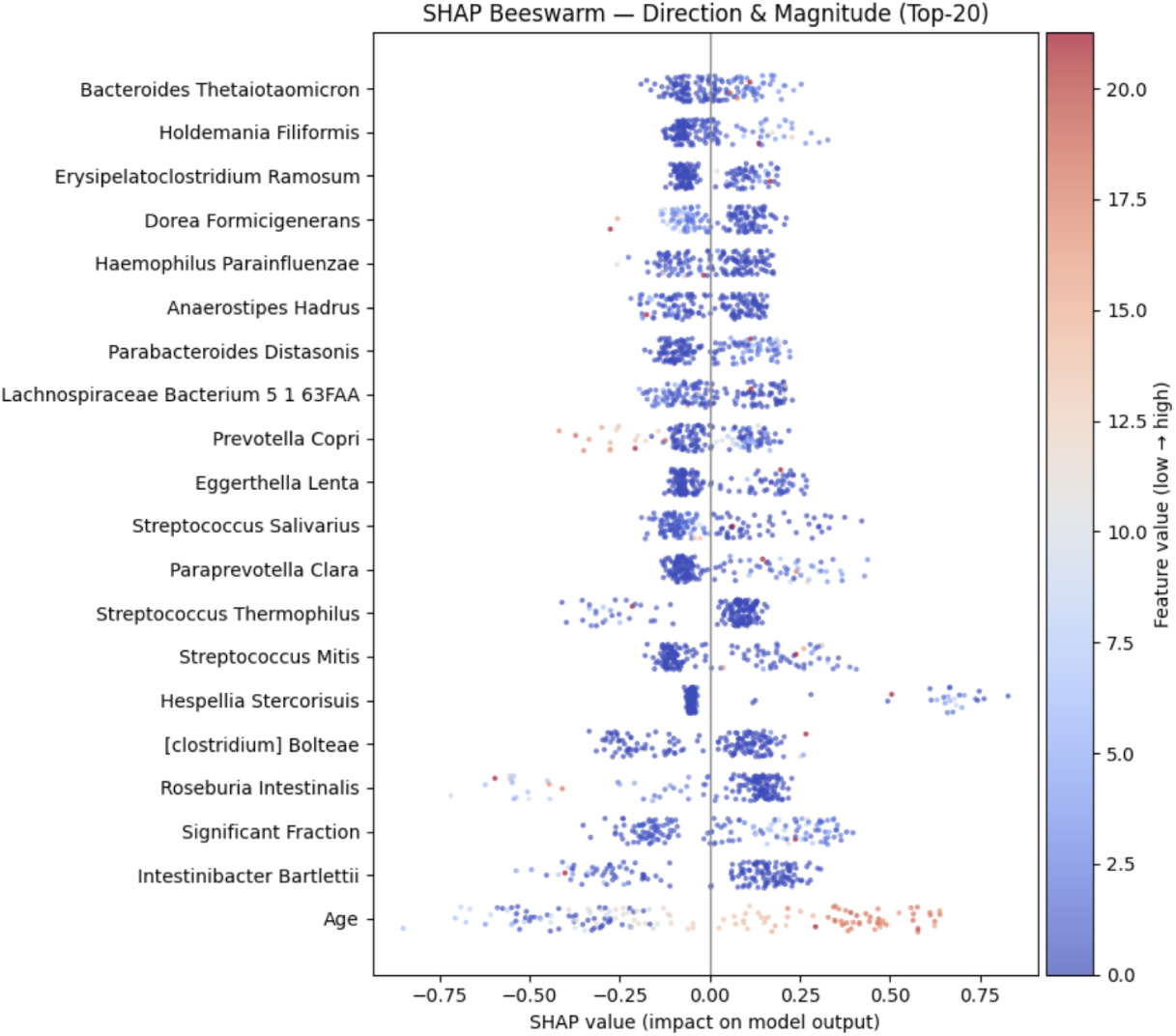
SHAP beeswarm plot across samples of the XGBoost model.

Since age is one of the factors, the range is much wider, showcasing a clear spread between lower and higher values; however, the bacterial species possess a shorter range of abundance values, depicted by the gradation of blue values. Many taxa show directionality consistent patterns, extending the biological plausibility found in the global feature importance plot. Nevertheless, certain bacteria show mixed directionality, in line with the idea that their contribution to either predicting a sample as healthy or diabetic depends on the context of other species or metadata specifications (i.e. age).

The stability of bacterial features selected using the SelectKBest method were validated in Figure 13, highlighting feature selection performed across 100 bootstraps. The original dataset samples were taken with replacement to create new datasets of equal sizes but varying compositions. All displayed taxa were selected in ≈0.63-0.71 bootstraps, highlighting the robustness and reproducibility of the bacterial features selected by the model. This demonstrates the biological validity of the features of these features by SelectKBest. Further, the absence of any single biomarker dominating with a near 100% stability highlights the multi-taxa considerations taken for the classification of any sample, justifying the biological grounding of the model. Similarly, the absence of very low-frequency features (<0.4) supports model stability and the effectiveness of feature filtering in eliminating non-significant features that would otherwise highly fluctuate between bootstraps.

**Figure 13:**
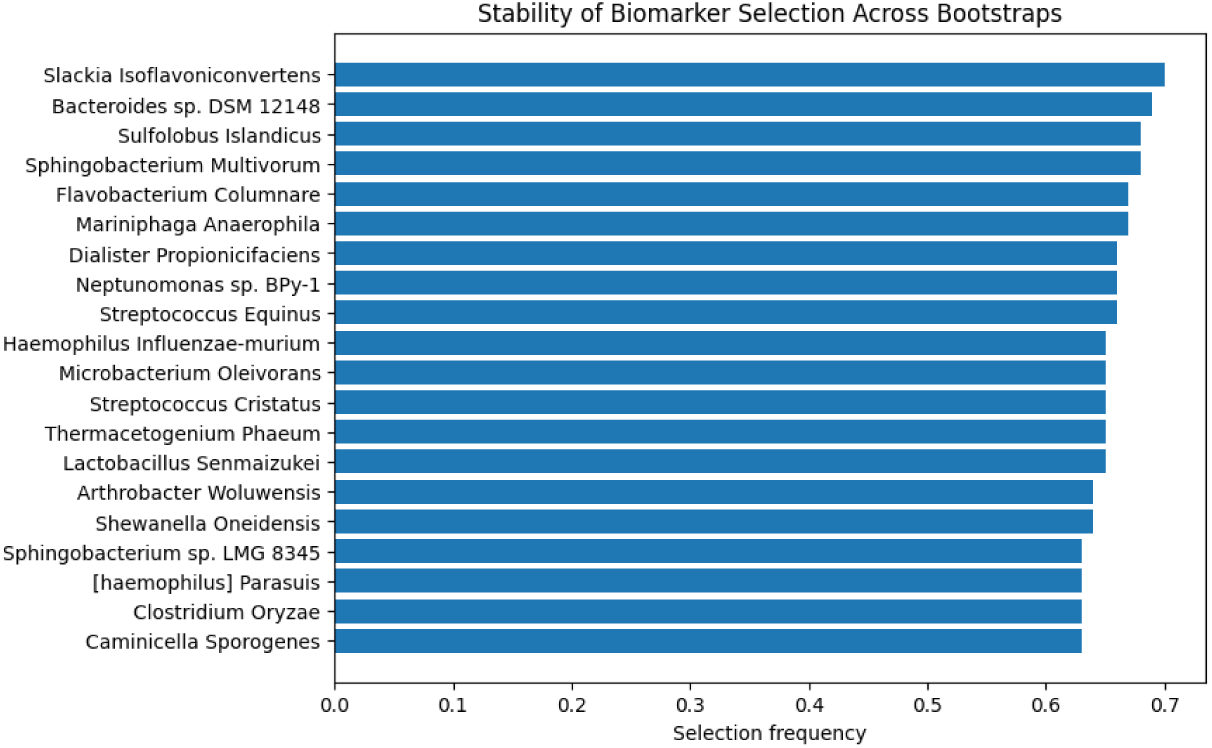
SHAP beeswarm plot across samples of the XGBoost model.

### 3.4 Necessity of Multivariate Modelling over Univariate Analyses

Volcano plots are employed to understand the differential expression of features present in a given dataset between the classes (in this case, diabetes and healthy control). The volcano plot (Figure 14) illustrates differential abundance of microbial species between diabetic and healthy cohorts. The x-axis represents the log-2 fold change, while the y-axis shows the -log10 (p-value). Species enriched in diabetic samples appear on the right, while those depleted appear on the left. The horizontal dashed line at -log10=1.30 indicates the significance threshold taken by the plot as p=0.05.

The absence of any points indicates that no single microbial species possesses both a strong differential abundance alongside a statistical significance between the two classes. Rather than suggesting a lack of microbiome variation between the two classes, this emphasizes the limitation of univariate statistical testing in capturing complex, interdependent microbial relationships. By demonstrating this insufficiency, the chart justifies the need for machine learning, which can integrate multiple features to analyse patterns and correlations between features, revealing underlying community-level patterns.

**Figure 14:**
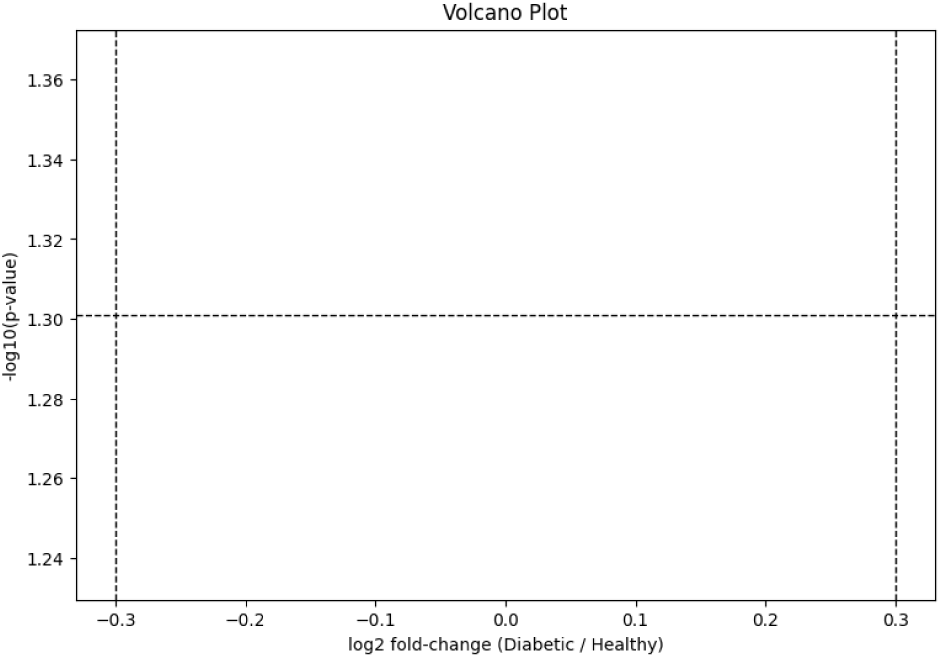
Volcano plot exhibiting the magnitude of and reliability of differential feature expression.

To illustrate the inter-feature relationships within the microbiome dataset, a correlational network map was created, as shown in Figure 15. Each microbial taxa is represented by a node, with the edges between the nodes depicting the correlations seen. Thicker edges represent stronger correlations, and groups of microbes seen in clusters are termed modules, acting as functional groups that co-occur. The presence of multiple sub-communities indicates microbial co-participation in pathways linked to diabetes status. The high interconnectivity reflects the co-variance of taxa, reinforcing the interdependent structure of the microbiome. Therefore, the presence of a structural and functional network demands the use of multivariate analysis and machine learning to capture network-level dependencies as compared to single-taxon statistical testing.

**Figure 15:**
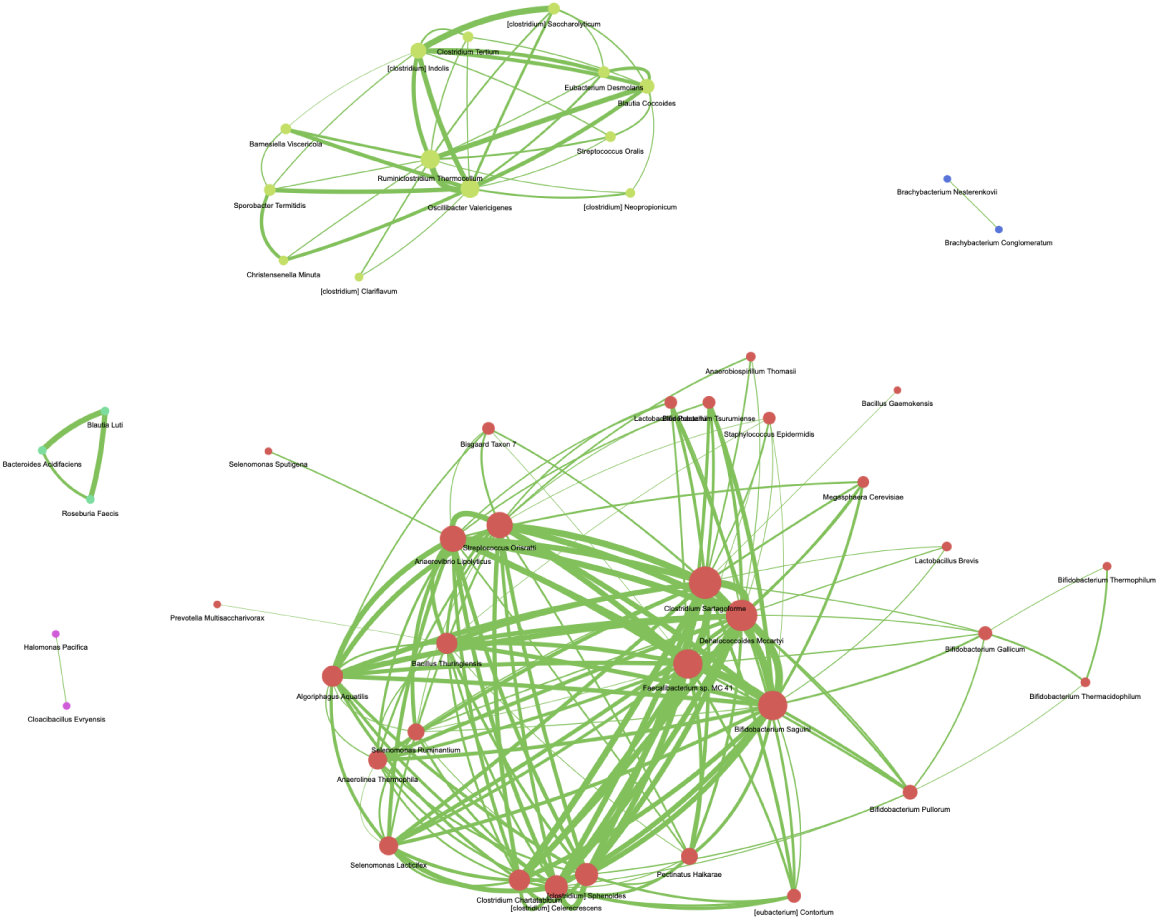
Correlational network map depicting relationships between bacterial species.

The above two graphs exemplify the need for machine learning to be employed in microbiome data for classification applications, as compared to traditional statistical approaches to such tasks.

### 3.5 Non-Linear Feature Interactions and Local Explanations

The SHAP force plot (Figure 16) illustrates how the model generates a single prediction. Those features moving to the right indicate their push towards the positive (diabetic) class, contrasting features to the left increasing the prediction probability of the negative (healthy) class. An E[f(X)] value of 0.055 represents the average classification over all samples (baseline model prediction). An f(x) value of 0.776 indicates the final model output as a confidence score of the prediction (in this case, diabetic given a positive value). The chart examines the key drivers influencing this decision, with where the value in the bar represents the magnitude of its SHAP contribution, and the y-axis values beside the feature indicate their abundance/quantity for the given sample. Collectively, these features shift the prediction toward the diabetic class, demonstrating how clinical and microbial factors interact to shape the model’s final classification.

**Figure 16:**
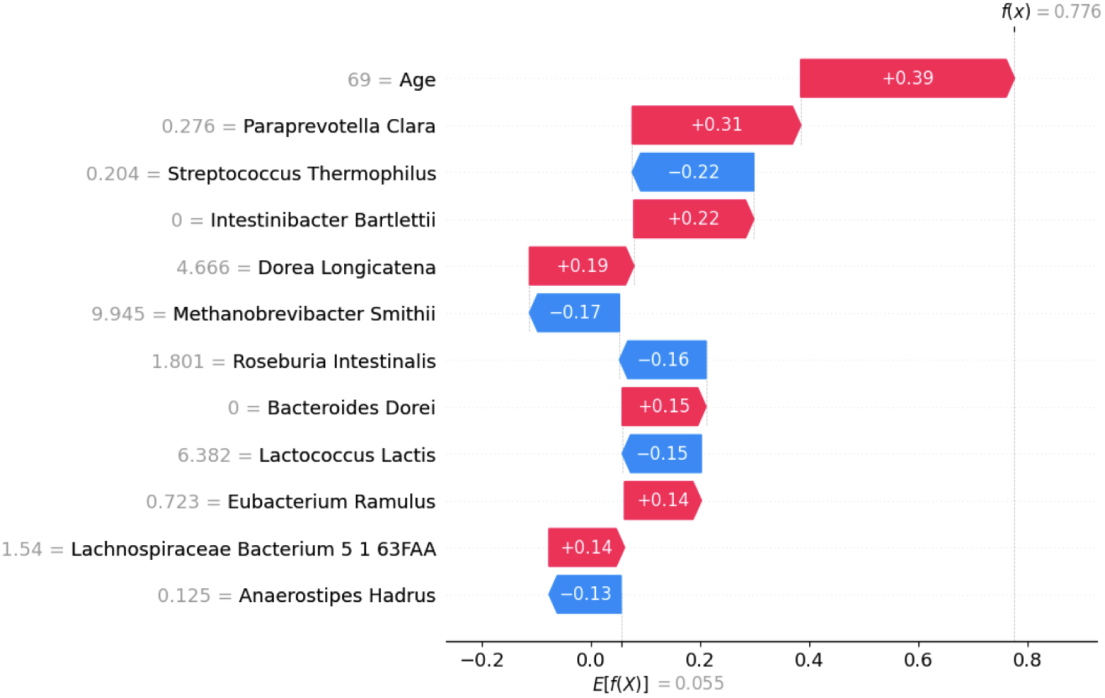
SHAP force plot for a given sample.

The SHAP decision plot (Figure 17) illustrates the contribution of multiple features on predictions made across samples. Unlike the force plot, which explains a single prediction, the decision plot aggregates feature contributions across multiple patients. This helps highlight those features pushing predictions up or down, and their consistency across samples. Age emerges as a consistent driver, where red (higher) feature values push the prediction to the right (diabetic class), highlighting the positive relationship between age and disease detection. Microbial taxa such as *Clostridium bolteae* are visible positive influencers of diabetes, where a higher abundance aligns with greater likelihood of a given patient having diabetes. Conversely, species like *Roseburia intestinalis* are negative contributors, indicating protective association as they push predictions towards the healthy class.

**Figure 17:**
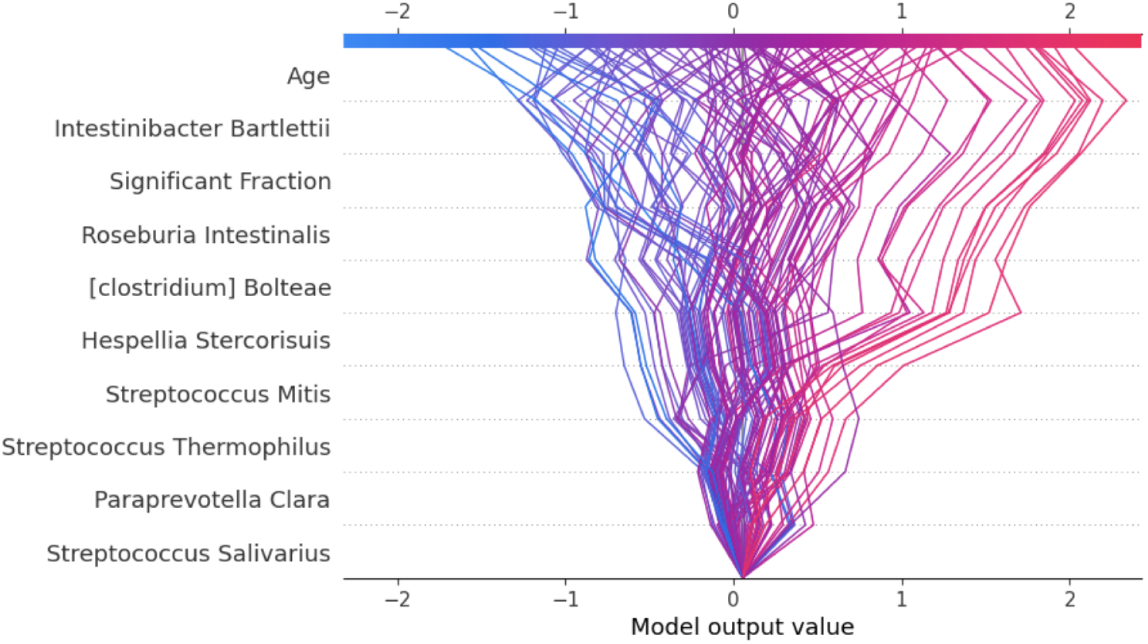
SHAP decision plot across multiple samples.

### 3.6 Biological Validation and Discovery of Potential Novel Biomarkers

In order to biologically validate the basis on which the research was crafted, namely the presence of varying abundances in gut microbiome species between diabetic (red) and healthy (blue) classes, violin plots were constructed as shown in Figure 17. Given the distinct abundance values between the two groups, a clear validation of gut biomarkers for diabetes is present, with either more or less enriched species in samples with the disease. Bidirectional shifts in abundance further strengthen the biological validity of the dataset, supporting a multi-feature approach over single-taxon analysis. Further, compared to the healthy violins, the diabetic violins are wider, indicating greater intra-group variance that highlights microbiome instability and dysbiosis, a hallmark of metabolic disorders like T2DM. Finally, significant overlaps in bacterial taxa (*Prevotella copri*, *Hespellia stercorisuis*, *Clostridium bolteae*, among others) between the global SHAP values (Figure 10) and top SelectKBest features represented in the violin plots (Figure 18) demonstrate the validity of the most important features chosen by two feature selection methods.

**Figure 18:**
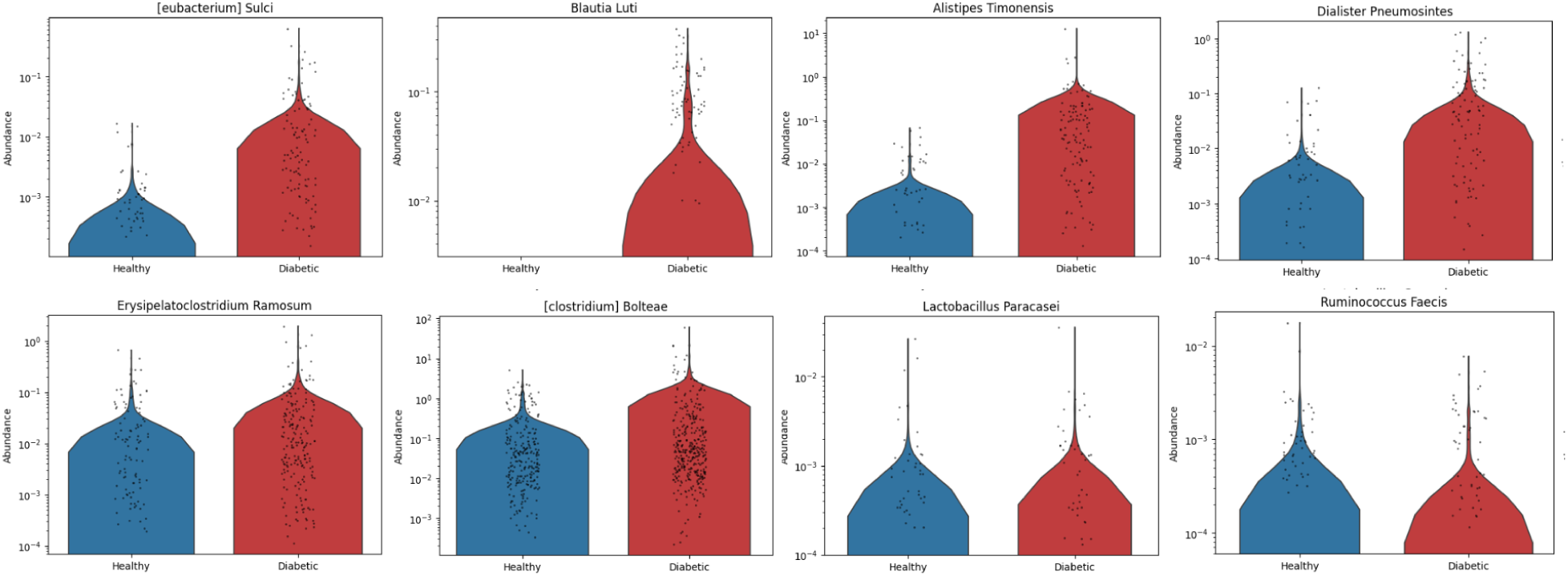
Violin plots of top features from SelectKBest, illustrating class-wise distribution differences.

The use of a heatmap, shown in Figure 19 served as another illustration to depict the differences between classes of the mean abundance of top biomarkers contributing to the model’s prediction.

**Figure 19:**
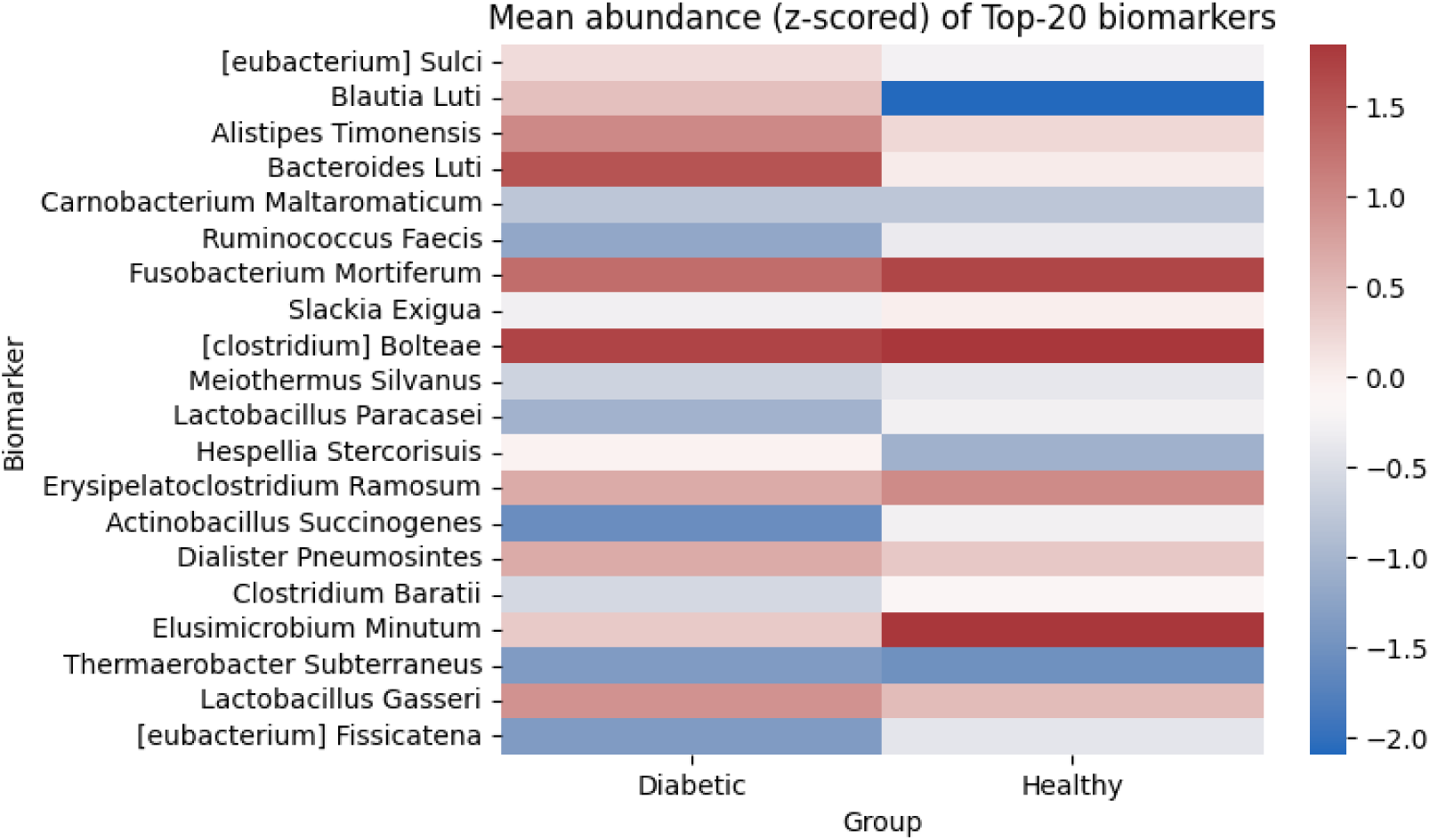
Heatmap of top features representing differences in mean abundances between classes.

The features represented in the violin plots and the SHAP analyses were compared with those identified gut biomarkers from the literature review. While most were bacterial species that were known, well-researched diabetic signatures, there were certain taxa that had not been documented as biomarkers of T2DM. Further research was conducted on such species, examining potential pathways and intermediary biological components that would correlate the given species with known T2DM processes. For instance, *Lactobacillus paracasei* is known as a probiotic in therapeutic contexts, though it has not been associated with diabetes. However, certain studies prove that in animal models, supplementation of the bacteria improved hyperglycemia, reducing inflammatory markers and also supporting an improved gut-barrier integrity^31,32^.

*Alistipes timonesis* is another taxa which, while correlated with gut dysbiosis overall, has not been classified as a T2DM marker. Its production of unique metabolites (such as sulfonolipids) tie it with diet-linked gut changes^33^; given diet is a strong risk factor of diabetes and high-fat/high-animal-based diets were associated with increased levels of *Alistipes*^34^, it can be hypothesized that the given species is a potential diabetes biomarker. The identified up-regulation in the diseased state, when further justified by the evident increase of the taxa’s abundance in the diabetic samples shown in the violin plots, indicates the plausibility of this gut microbe as a possible diabetic indicator. Another species with the genus *Eubacterium* contains multiple species that produce butyrate^35^. Given butyrate’s known role as a metabolite linked to gut integrity and insulin sensitivity^36^, thereby indicating that the bacteria’s presence in the butyrate-producer network could correlate it with T2DM risk.

### 3.7 Synbiotic-Mediated Pathway Regulation

Individual patient compositions led to insights on potential synbiotic formulations that could be administered to upregulate or downregulate bacterial species. Key biomarkers deviating from healthy thresholds were specifically targeted using synbiotics, such as *Lactobacillus rhamnosus GG* and *d-tagatose* to upregulate *Lactobacillus*^37^, as shown in Figure 20. Species were ranked by importance and degree of deviation from the healthy median. For each key feature identified, corresponding synbiotic candidates were recommended. Patient-specific taxa show significant individual variation, thereby supporting personalised synbiotic design.

**Figure 20:**
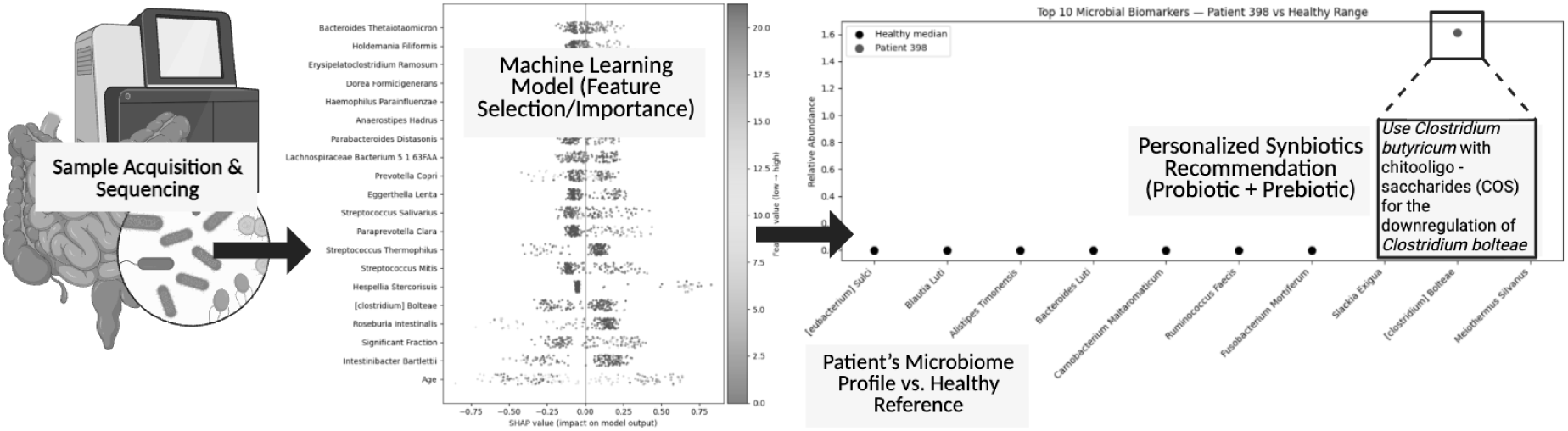
Individual Microbiome Profiling for the development of Personalized Synbiotic composition.

Expanding the current repository of experimentally validated synbiotics (Table 6) could enhance intervention design by broadening the range of targetable biomarkers. Further experimental validation of untested synbiotics is essential to ensure clinical reliability. Nonetheless, synbiotics remain a promising route for modulating influential taxa, enabling personalized and targeted treatment strategies.

**Table 6:** Excerpt of Synbiotic Intervention Data Used in Post-Patient Analysis.

| Microbe Name | Substance Name | Substance-Category | Substance-subCategory | Microbe Change | Condition/Disease | Reference_ID |
| --- | --- | --- | --- | --- | --- | --- |
| *Bifidobacterium longum* | Inulin + B. longum | Synbiotic | Prebiotic + Probiotic | Increase | Type 2 Diabetes | PMID: 31456712 |
| *Lactobacillus rhamnosus* | GOS + L. rhamnosus | Synbiotic | Prebiotic + Probiotic | Increase | Obesity/T2D | PMID: 30011245 |
| *Faecalibacterium prausnitzii* | FOS + B. adolescentis | Synbiotic | Prebiotic + Probiotic | Increase | Metabolic Syndrome | PMID: 28945121 |
| *Akkermansia muciniphila* | Polyphenol mix + L. plantarum | Synbiotic | Plant Prebiotic + Probiotic | Increase | Insulin Resistance | PMID: 29123456 |
| *Roseburia intestinalis* | Resistant starch + L. casei | Synbiotic | Prebiotic + Probiotic | Increase | Gut Dysbiosis | PMID: 32098712 |

### 3.8 FMT Donor Predictive Scoring

According to the methodology detailed in Section 2.4, donor-recipient rankings were generated for all the diabetic patients, with the top 20 results summarised in Table 7. For a given representative recipient, the corresponding bar chart (Figure 21) visualises these rankings. The FMT rankings and match-scores were calculated for 50 different recipients (diabetic patients), across which donor match scores exhibited a high range. Top-ranking donors fell between the ≈93.9-94.9 band as shown in the bar plot, highlighting the strong compatibility of a set of donors for certain recipients. The distribution of donor scores highlighted the influence of variations in the magnitude of a certain microbiome community’s presence on their suitability to the recipient. While generalized across the 50 recipients, such results highlight the existence of multiple possible donor matches for a given diabetic patient undergoing FMT. Further, variability among donor ranks between different recipients highlights the importance of personalised, recipient-specific donor-matching as compared to a universal preference for any given donor.

**Table 7:**
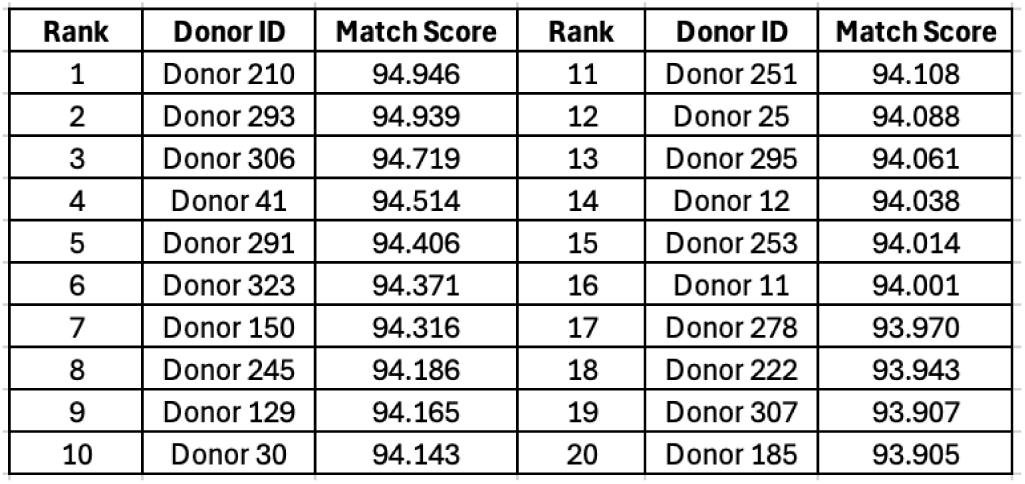
FMT donor–recipient match scores and rankings for a given diabetic recipient.

| Rank | Donor ID | Match Score | Rank | Donor ID | Match Score |
| --- | --- | --- | --- | --- | --- |
| 1 | Donor 210 | 94.946 | 11 | Donor 251 | 94.108 |
| 2 | Donor 293 | 94.939 | 12 | Donor 25 | 94.088 |
| 3 | Donor 306 | 94.719 | 13 | Donor 295 | 94.061 |
| 4 | Donor 41 | 94.514 | 14 | Donor 12 | 94.038 |
| 5 | Donor 291 | 94.406 | 15 | Donor 253 | 94.014 |
| 6 | Donor 323 | 94.371 | 16 | Donor 11 | 94.001 |
| 7 | Donor 150 | 94.316 | 17 | Donor 278 | 93.970 |
| 8 | Donor 245 | 94.186 | 18 | Donor 222 | 93.943 |
| 9 | Donor 129 | 94.165 | 19 | Donor 307 | 93.907 |
| 10 | Donor 30 | 94.143 | 20 | Donor 185 | 93.905 |

**Figure 21:**
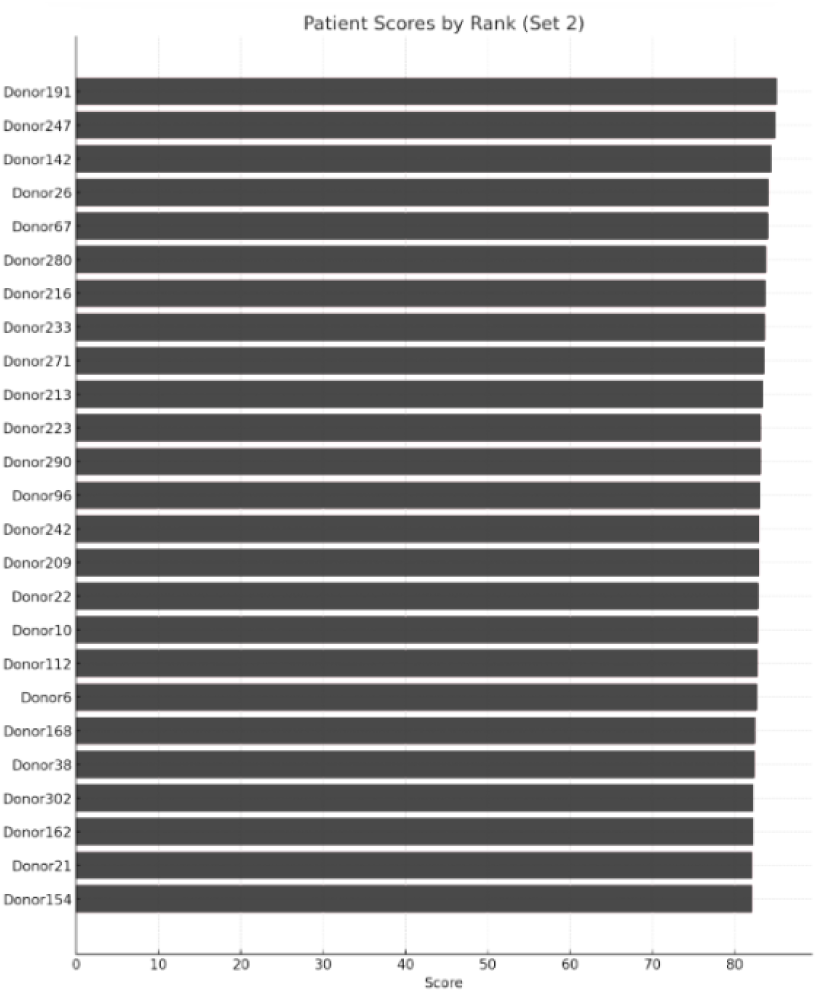
Visualising donor FMT match scores and rankings for a diabetic recipient.

By compiling the results of 10 of the 50 recipients for visualisation purposes, a heatmap was constructed, depicted in Figure 22. Of greatest significance is the finding that donor-recipient compatibility patterns are heterogeneous, shown in the variations in efficacy of a given donor for different recipients. The presence of certain donors possessing high matching scores across multiple recipients does hint at a potential ‘broad spectrum donor’ status; however, personalised donor selection is in almost every case more effective to the universal selection of any one given donor across recipients. The variation in range and distribution of donor compatibility scores across recipients indicate some diabetic patients could be easier to match with healthy donors, while others require a larger pool for finding effective matches. Finally, clustering within the heatmap indicates groups of donors sharing microbiome features that confer similar match profiles.

**Figure 22:**
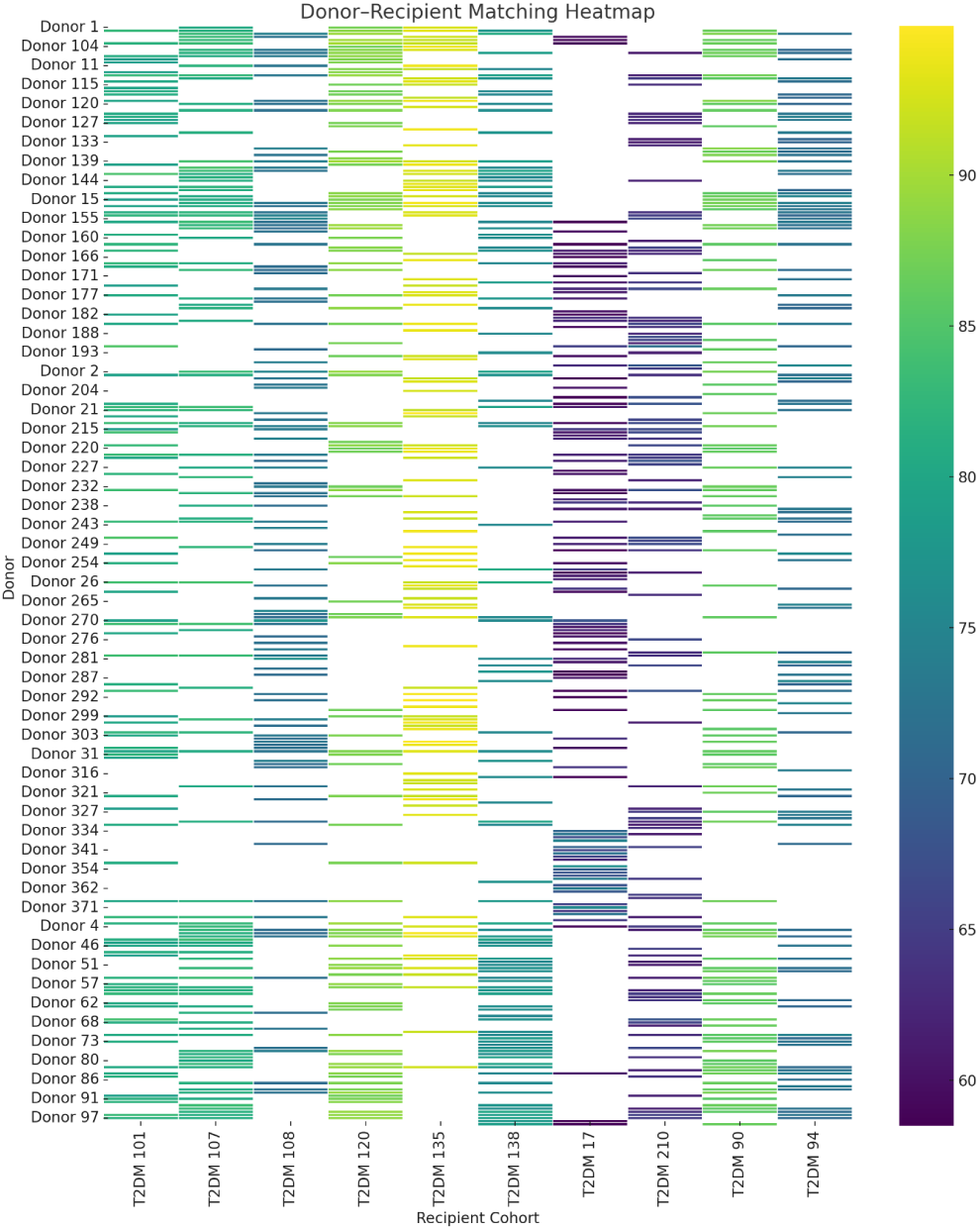
Heatmap illustrating donor–recipient match scores for FMT.

In order to map trends within and across recipients, boxplots were constructed (Figure 23). These charts further extrapolated the idea of inter-recipient variability, wherein certain diabetic patients are easily matched by the set of donor samples taken while others possessing lower match scores are harder. However, the presence of higher outliers even for those recipients with lower average scores (such as ‘T2DM 17’, ‘T2DM 210’, ‘T2DM 108’) indicate that even such patients have donors that are highly compatible, despite being isolated. In comparison, certain recipients (including ‘T2DM 135’, ‘T2DM 120’, ‘T2DM 90’) demonstrate high score ranges with low variance, implying their greater matchability for the given healthy sample group chosen. Recipients with lower inter-quartile ranges suggest more robust donor predictions, while wider intervals indicate high donor-specific performance fluctuations. The construction of such a box plot exemplifies the importance of the continuous nature of donor-recipient match data, as compared to a binary classification that lacks the sophistication of a computed score range.

**Figure 23:**
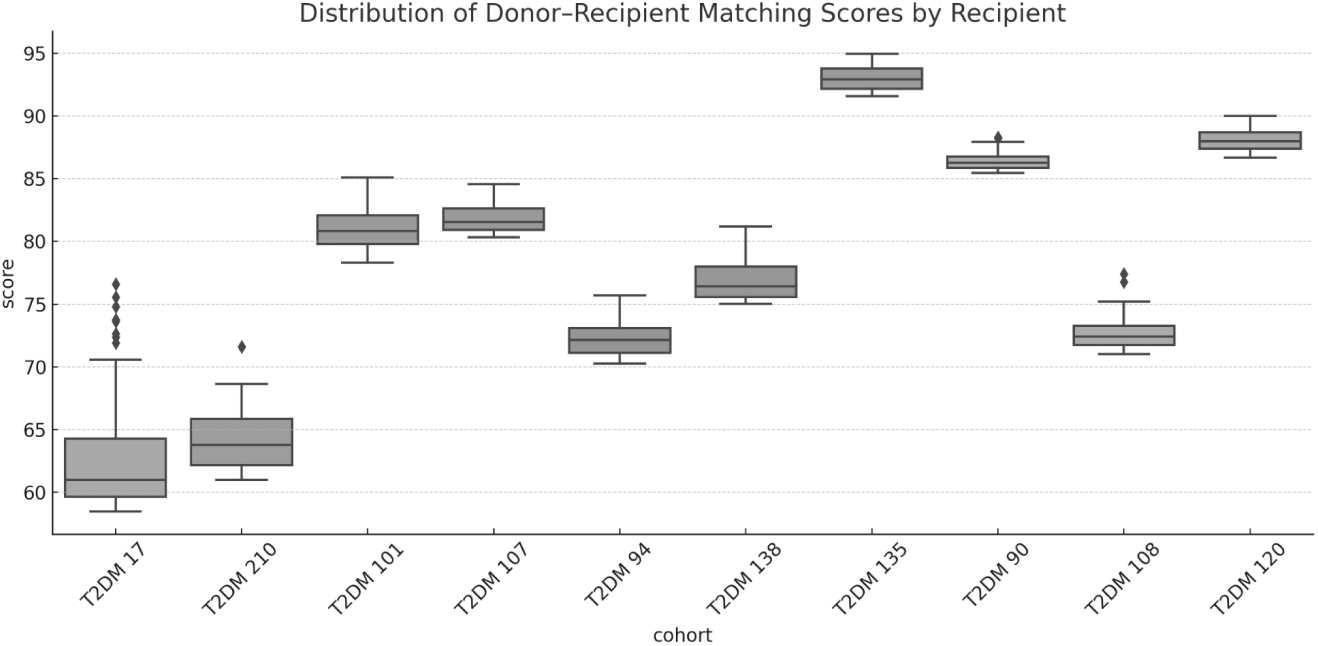
Highlighting trends in donor–recipient matching scores for 10 sample patients.

## 4. Conclusion

The study employed a machine learning model to classify patients between T2DM and healthy classes based on their gut microbiome readings, aiding improved disease diagnosis and targeted intervention. Such an approach transcends traditional statistical analyses that often use univariate comparisons between groups for predictions, instead employing a multi-variate approach better aligned with interdependencies between biological features. When seen in the context of the nature of microbiome data (with a 95.8% sparsity in the training set employed for this study), the achieved 80.00% test accuracy highlights the immense potency of such computational techniques on enabling early detection and intervention. Microbiome-driven disease diagnostics can complement standard glucose and metabolite tests, especially due to its ability to offer a scalable, non-invasive diagnostic that complements precision medicine and personalised therapeutic interventions.

An outcome of the study was the high robustness of ML-models in maintaining biological integrity and remaining reliable to the pathophysiological nature of a given disease. The construction of SHAP analyses, correlational network maps and violin plots confirm the top predictive taxa used in feature selection methods are highly representative of documented species. The employment of feature analysis further supported the identification of novel or under-reported taxa that had a highly plausible connection to T2DM processes through various pathways and metabolic intermediaries. Such results highlight the study’s informed findings of potential biomarkers that require further research to expand the existing literature on the topic.

Given only ≈4% of the data used contained non-zero values, the ability for the model to generalise findings towards an 80% testing accuracy, precision and recall, demonstrate the strong robustness of the created model to noise and class imbalance. Especially when put in context of past efforts to train models on microbiome data, the crafted XGBoost model’s performance metrics show significant improvement, especially when applied across studies for generalization analysis. The greater success achieved by the boosting model can largely be attributed to biologically-informed feature selection (GBM and SelectKBest) and regularized designs to minimize overfitting.

The integration of two interventions, FMT and synbiotics, further emphasises the translational value of the model in supporting therapeutic modelling. The pipeline’s ability to provide personalised microbiome restoration strategies by pinpointing up/down-regulation highlights the widespread potential of such computational methods. Using precision synbiotic design and donor-recipient matching algorithms, the system developed acts as a closed-loop diagnostic-intervention cycle enabling prediction, validation and personalisation.

The largest constraint in deploying such models is the dearth of open-source disease-specific microbiome data that ensure standardization of data collection and demographic (metadata) diversity and availability. Future research studies that produce data must integrate host metadata consistently, prevent intra-study sample variation for key criteria (such as the presence of other chronic diseases or illnesses), and expand sample-size. Such improvements can enable the use of more sophisticated models to improve inter-study classification validity and prediction accuracy.

Future directions for this research include: a) assimilating datasets with larger sample sizes for improved feature-to-sample ratio, enhancing performance; b) testing model robustness through mimic disease incorporation (obesity, T1DM, other related diseases); c) integrated intermediary stages (prediabetic) between healthy and diabetic for early intervention; d) expanding personalised therapeutic strategies, such as testing efficacy of diet interventions; e) collecting human gut samples from patients belonging to the two cohorts, to conduct a double-blind study on disease status prediction.

By demonstrating the ability for machine learning to classify diabetic patients with high accuracy even in ultra-sparse microbiome environments, staying rooted in biologically-grounded networks and co-occurrences, this work marks a pivotal step toward data-driven, personalised diabetic diagnostics and interventions.

## Data Availability

All data produced in the present study are available upon reasonable request to the authors

